# Elevated plasma Complement Factor H Regulating Protein 5 is associated with venous thromboembolism and COVID-19 severity

**DOI:** 10.1101/2022.04.20.22274046

**Authors:** Laura Sanchez-Rivera, Maria Jesus Iglesias, Manal Ibrahim-Kosta, Julia Barbara Kral-Pointner, Sebastian Havervall, Louisa Goumidi, Maria Farm, Gaëlle Munsch, Marine Germain, Philip Smith, Mun-Gwan Hong, Pierre Suchon, Clément Naudin, Anne Boland, David M Smadja, Margareta Holmström, Maria Magnusson, Angela Silveira, Mathias Uhlén, Thomas Renné, Angel Martinez-Perez, Joseph Emmerich, Jean-Francois Deleuze, Jovan Antovic, Alice Assinger, Jose Manuel Soria Fernandez, Charlotte Thålin, Jochen M Schwenk, Juan Carlos Souto Andres, Pierre-Emmanuel Morange, Lynn Marie Butler, David-Alexandre Trégouët, Jacob Odeberg

**Author notes:** Address correspondence to: Jacob Odeberg Science for Life Laboratory, Department of Protein Science, CBH, KTH Royal Institute of Technology, SE-171 21 Stockholm, Sweden or David-Alexandre Trégouët University of Bordeaux, Inserm, Bordeaux Population Health Research Center, UMR 1219, team ELEANOR, Bordeaux, France. **One sentence summary:** Elevated CFHR5 plasma concentration is associated with diagnosis and risk of venous thromboembolism, and with short-term prognosis in hospitalised COVID-19 patients. Equal contribution (Laura Sanchez-Rivera, Maria Jesus Iglesias). Equal contribution (Pierre-Emmanuel Morange, Lynn Marie Butler, David-Alexandre Trégouët, Jacob Odeberg).

## Abstract

Venous thromboembolism (VTE), comprising both deep vein thrombosis (DVT) and pulmonary embolism (PE) is a common, multi-causal disease with potentially serious short- and long-term complications. In clinical practice, there is a need for improved plasma biomarker-based tools for VTE diagnosis and risk prediction. We used multiplex proteomics profiling to screen plasma from patients with suspected acute VTE, and a case-control study of patients followed up after ending anticoagulant treatment for a first VTE. With replication in 5 independent studies, together totalling 1137 patients and 1272 controls, we identify Complement Factor H Related Protein (CFHR5), a regulator of the alternative pathway of complement activation, as a novel VTE associated plasma biomarker. Using GWAS analysis of 2967 individuals we identified a genome-wide significant pQTL signal on chr1q31.3 associated with CFHR5 levels. We showed that higher CFHR5 levels are associated with increased thrombin generation in patient plasma and that recombinant CFHR5 enhances platelet activation *in vitro*. Thrombotic complications are a frequent feature of COVID-19; in hospitalised patients we found CFHR5 levels at baseline were associated with short-time prognosis of disease severity, defined as maximum level of respiratory support needed during hospital stay. Our results indicate a clinically important role for regulation of the alternative pathway of complement activation in the pathogenesis of VTE and pulmonary complications in acute COVID-19. Thus, CFHR5 is a potential diagnostic and/or risk predictive plasma biomarker reflecting underlying pathology in VTE and acute COVID-19.

## INTRODUCTION

Venous thromboembolism (VTE), comprising both pulmonary embolism (PE) and deep vein thrombosis (DVT) is a common, multi-causal disease with serious short and long-term complications. VTE has a high mortality rate in the first year, especially within the first 30 days (∼30% for PE) and a high risk of recurrence, with a cumulative incidence rate of 25% within 10 years [1–4]. VTE diagnosis is challenging, as the predisposing common risk factors and clinical presentation can be consistent with multiple other conditions, particularly in the case of PE. Current VTE diagnostic work-up includes assessment of clinical probability, using clinical decision rules, e.g., the Well’s score, in combination with elevated plasma D-dimer levels [5, 6]. However, D-dimer, a clot breakdown product, can be elevated in several other non-VTE conditions (e.g., inflammation, surgery, cancer). Thus, its usefulness is limited to ruling out VTE in low probability cases. In medium and high probability cases, diagnostic imaging, i.e., compression ultrasound (CUS) on suspicion of DVT, or computed tomography pulmonary angiogram (CTPA) on suspicion of PE, is necessary to exclude or confirm diagnosis. These tests may not be readily available in the acute setting, resulting in delayed or incorrect diagnosis. Today, less than 20% of CTPAs and CUS confirm suspected PE [7–9] or DVT [10], respectively. Specific biomarker-based tools in the diagnostic workup could reduce unnecessary imaging, save resources and, in the case of CTPA, reduce exposure to radiation and contrast agent. Several studies have proposed VTE biomarker candidates (e.g., p-selectin, microvesicles) [11], but none have reached clinical implementation. There is also a need for improved tools and plasma biomarkers for risk prediction in VTE. Risk scores based on clinical risk factors and D-dimer levels have been developed for recurrence prediction [12–18], but none are routinely integrated into clinical practice. In addition to the genetic variants currently used in clinical assessment of hereditary thrombophilia (e.g., Factor V Leiden, prothrombin mutation) there are other more recently discovered common gene variants that contribute to VTE risk [19–21]. However, even when these are also incorporated into risk scores, they still lack sufficient precision for VTE prediction on an individual basis [22, 23]. This likely reflects the interplay between transient and sustained risk factors in disease development, including acquired risk factors, genetics, and environmental exposures [24, 25].

VTE is a disease of the intravascular compartment and thus analysis of the blood proteome has the potential to capture the resulting effects of combined genetic, epi-genetic, and environmental contributors to risk variation. Large scale plasma proteomics screening allows for discovery of novel protein biomarkers with potential clinical utility for diagnosis and/or prediction. Such studies could also reveal biological pathways involved in VTE pathogenesis, for further functional studies to identify targets for tailored treatment. So far, a handful of plasma proteomics studies of VTE have been reported, presenting novel candidates associated with increased risk of VTE [26–30], including our own; the first affinity proteomics case-control study of patients followed up post treatment after a first VTE, reporting PDGFB as a novel VTE associated biomarker [27]. High throughput affinity proteomics studies have increased the capacity for discovery screening and identification of novel associations, as now it is possible to screen for hundreds or thousands of proteins in small plasma volumes [31]. Orthogonal verification is essential to confirm results generated using these technologies but is frequently absent from such studies.

Here, we aimed to identify novel biomarkers associated with acute VTE with a potential link to underlying VTE pathogenesis. We identified complement factor H-related protein 5 (CFHR5), a regulator of the alternative pathway of complement activation, as a novel VTE associated plasma biomarker. Furthermore, we found that CFHR5 plasma levels are associated with short term prognosis in acute COVID-19, a disease where thrombosis formation is central to pathology. Our study suggests that CFHR5 could be involved in the underlying pathogenesis of VTE and acute COVID-19, and that it is a potential clinical biomarker for thrombotic disease diagnosis and/or risk prediction.

## RESULTS

### Affinity plasma proteomics identifies candidate proteins associated with acute VTE

To identify plasma biomarkers for VTE we analysed samples collected as part of our *Venous thromboEmbolism BIOmarker Study* (*VEBIOS*) [27]. *VEBIOS* comprises two study arms; a prospective cohort of patients sampled at the Emergency Room (ER), Karolinska University Hospital, Sweden (*VEBIOS ER*) and a case/control study with patients sampled at an outpatient coagulation clinic after discontinuation of anticoagulant treatment after a first VTE (*VEBIOS Coagulation*). The discovery cohort, *VEBIOS ER* (Figure 1A), consisted of patients (n= 147) admitted to the ER with the suspicion of DVT in the lower limbs and/or PE. Following admission, two whole blood samples were collected from participants, into citrate and EDTA anticoagulant. Patient samples were classified as controls (n=96), when a VTE diagnosis was excluded by diagnostic imaging, and/or Well’s clinical criteria with a normal D-dimer test, or cases (n=51) when VTE was confirmed by diagnostic imaging and anticoagulant treatment was initiated. A nested case/control sample set of 48 cases and 48 matched controls were selected for plasma protein analysis (Table 1). Target candidates for measurement were selected as previously described [27], based on: (i) indications from the literature, in house data or public repositories of a probable or plausible link to arterial or venous thrombosis (e.g., prior evidence of association with thrombosis or intermediate traits, or known involvement in biological pathways of relevance), including 124 that we predicted to have endothelial enriched expression [32], and (iii) the availability of target specific antibodies in the Human Protein Atlas (HPA). A total of 758 HPA antibodies, targeting 408 candidate proteins, were selected for incorporation into a single-binder suspension bead array (Figure S1 A and Table S1, Tab_1), which was used to analyse plasma generated from the blood samples collected into citrate anticoagulant. The signal generated by antibody HPA059937, raised against the protein target sulfatase 1 (SULF1), was most strongly associated with VTE (p<8.34E-06) (Figure 1B, green point), with higher relative plasma levels in cases vs. controls (Figure 1D.i). Signals generated by a further seven antibodies were also associated with VTE (p<0.01) (Table S1, Tab_1). Protein signatures in plasma can be differently affected by the sample matrix; anticoagulants can inhibit specific proteases, influence soluble protein interactions, and modify analyte stability. Thus, the anticoagulant type used has potential consequences for biomarker identification [33]. We therefore replicated the *VEBIOS ER* discovery screen in EDTA samples drawn in parallel from the same patients (Figure 1C). Of the eight antibodies that produced signals associated (p<0.01) with VTE in the citrate samples (Table S1, Tab_1), four were replicated in the EDTA samples (Figure 1B and C): HPA059937 (predicted target SULF1, green point), HPA044659 (predicted target Leukocyte surface antigen CD47 [CD47], blue point), HPA003042 (predicted target Adenosine receptor A2a [ADORA2A], orange point) and HPA002655 (predicted target P-Selectin [SELP], red point) (Table S1, Tab_1). In both anticoagulants, all four target candidates were elevated in cases vs. controls (Figure 1D and E, i-iv). In all subsequent experiments, citrated blood was used in the analysis, on the basis that it is the more commonly used sample anticoagulant in the clinical setting for coagulation analyses.

**Figure 1:**
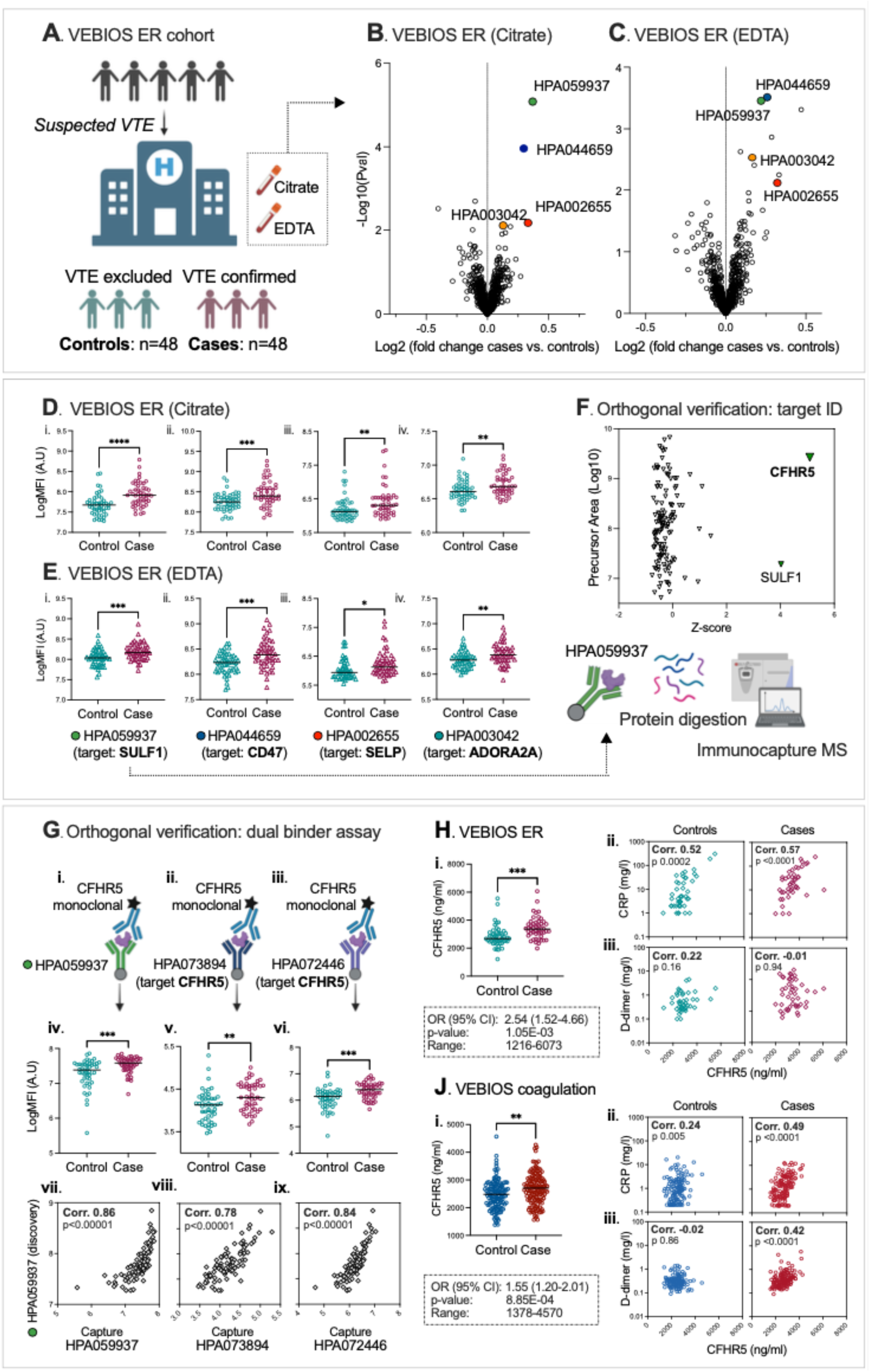
Plasma proteomics profiling identifies CFHR5 as associated with VTE. (**A**) Overview of the VEBIOS ER discovery cohort. 758 HPA antibodies, targeting 408 candidate proteins, were used to analyse plasma samples using affinity proteomics. Log fold changes in antibody MFI (mean fluorescent intensity) signal were calculated between VTE cases and controls in (**B**) citrate or (**C**) EDTA anticoagulated plasma; coloured circles indicate antibodies that generated signals significantly associated with VTE in both. MFI signals generated by these antibodies for controls and cases in (**D**) citrate plasma and (**E**) EDTA plasma. (**F**) Immunocapture-mass spectrometry identification of protein targets of HPA059937. (**G**) Dual binder assays were developed using an anti-CFHR5 detection antibody, combined with (**i**) HPA059937 (raised against SULF1) (**ii**), anti-CFHR5 HPA073894 or (**iii**) anti-CFHR5 HPA072446 as capture antibodies. CFHR5 levels in the citrated plasma samples were re-analysed, using the respective dual binder assays, to determine (**vii-ix**) levels (MFI) in controls vs. cases and (**vii-ix**) the correlation between the signal and those generated by the original single binder assay using HPA059937. Dual binder assay using capture antibody HPA072446 with a recombinant protein standard was used for absolute quantification of CFHR5 in samples from: (**H**) *VEBIOS ER* and (**J**) *VEBIOS Coagulation*. CFHR5 concentration was (**i**) measured in controls and cases, with associated OR (odds ratio per 1 standard deviation increase) or (**ii**) used to determine the correlation with C-reactive protein (CRP), or (iii) d-dimer concentration. ***p<0.0001, ***p<0.001, **p<0.01.

**Table 1.**
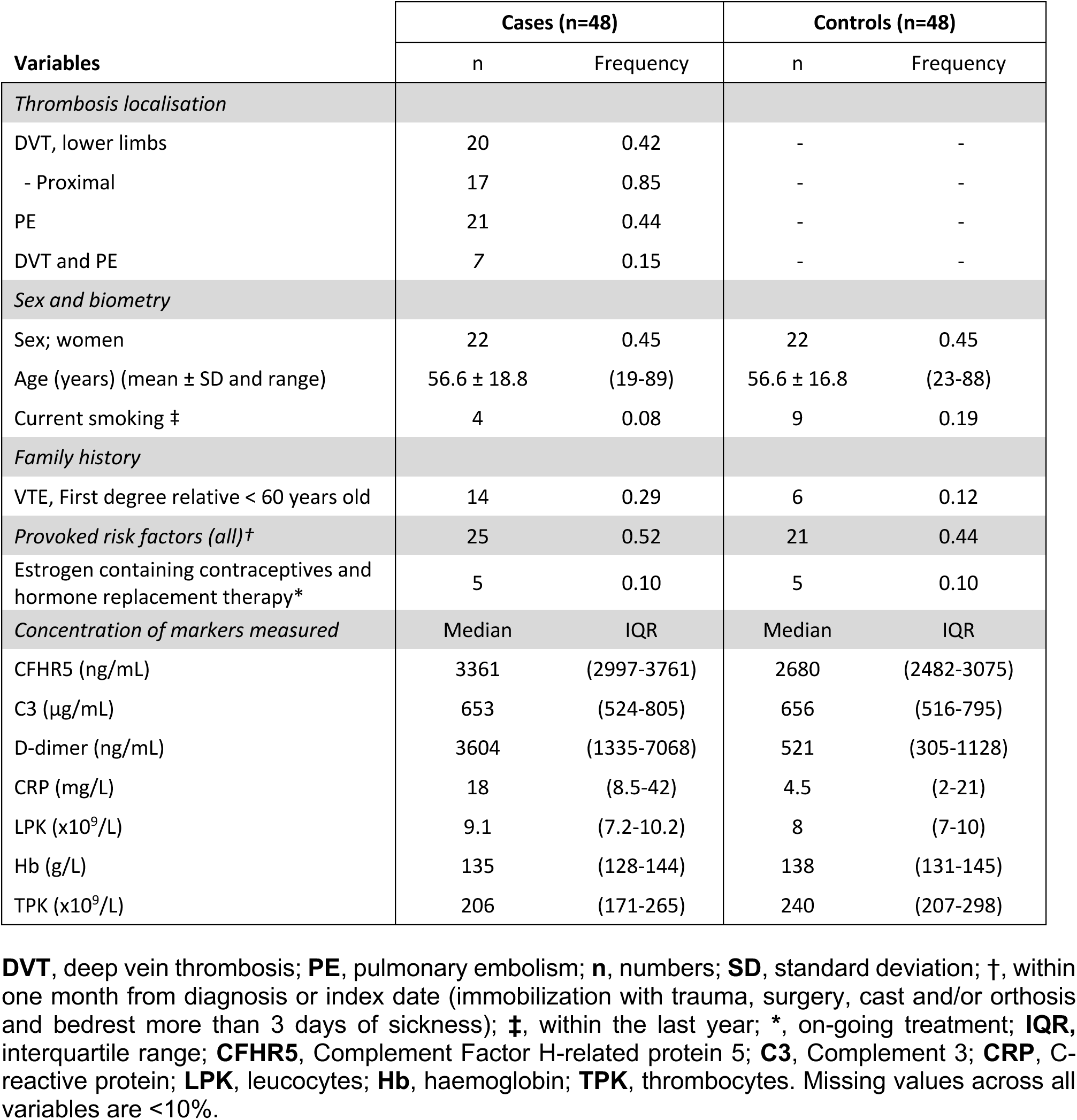
Clinical characteristics of the VEBIOS ER sample set.

Previously, in the *VEBIOS Coagulation* study (n=177) [27], we identified 29 protein candidates in plasma that were associated with prior VTE. This study was composed of patients sampled 1-6 months after discontinuation of anticoagulation treatment (duration 6-12 months) following a first time VTE, or matched controls. Of the four antibodies that generated signals associated with acute VTE in citrate *and* EDTA plasma in *VEBIOS ER* (Figure 1B and C, marked with coloured circles), only HPA059937 (predicted target SULF1), produced a signal associated with prior VTE in the *VEBIOS Coagulation* study [27]. As our aim was to identify biomarkers associated with acute VTE that were potentially linked to the underlying disease pathogenesis, we prioritised this candidate on the basis that higher plasma concentrations observed in individuals with a documented increased risk of VTE (e.g., previous VTE) in *VEBIOS Coagulation* study indicated that it could also represent a constitutive and/or persistent risk factor.

### Complement Factor H Related protein 5 (CFRH5) is associated with VTE

The antibodies used in the single-binder suspension bead arrays passed quality control for antigen binding specificity (see www.proteinatlas.org/), but selective binding to the target protein in context of the complex matrix of plasma requires verification, as antibody specificity and reliability can be a problematic issue [34–36] (Figure S1 C-F). To verify which protein(s) were captured by HPA059937 (predicted target SULF1) *w*e performed immunocapture-mass spectrometry (IC-MS). Two proteins were bound to HPA059937 with a z-score>3 in triplicate experiments; the predicted target, SULF1 (z-score = 4.02, with 1 Peptide Spectrum Match [PSM]), and Complement Factor H Related protein 5 (CFHR5) (z-score=5.09, with ≥21 PSM) (Figure 1F and Table S1, Tab_2). These data indicate that CFHR5 was the predominant protein captured by HPA059937 in plasma, whilst not ruling out concurrent binding of SULF1. High levels of CFHR5 have been detected in plasma by mass spectrometry (MS) [37], but SULF1 (a protein predicted to be localised on the cell surface) is below the MS detection threshold [38]. To further verify CFHR5 binding specificity of HPA059937, we developed three dual binder assays (Figure S1B and Figure 1G), all with a commercial monoclonal antibody against CFHR5 as a detection antibody, combined with either: original antibody HPA059937 (Figure 1G.i), or one of two independent antibodies raised in house against CFHR5; HPA073894 (Figure 1G.ii) or HPA072446 (Figure 1G.iii), as bead coupled capture antibodies. When used to re-analyse the *VEBIOS ER* samples; all three assays consistently detected a higher level of target protein in cases vs. controls (Figure 1G iv-vi, p=0.0001, 0.0021, 0.0006, respectively). Furthermore, MFI values from all three strongly correlated with those generated by HPA059937 in the *VEBIOS ER* discovery screen (Figure 1G.vii-ix) (Spearman correlation 0.86, 0.78 and 0.84, respectively [all p<0.00001]). We made five dual binder assays that targeted SULF1, but none gave a quantitative signal in a plasma dilution series, or buffer containing a dilution series of recombinant SULF1 protein. These data are consistent with CFHR5, as opposed to SULF1, being the target protein associated with VTE in the *VEBIOS ER* discovery screen.

### CFHR5 is associated with VTE independent of D-dimer or CRP

The CFHR5 dual binder assay using capture antibody HPA072446 (Figure 1G.iii), together with recombinant CFHR5 protein standard, was used for absolute quantification of CFHR5 in *VEBIOS ER* and an extended sample set of the *VEBIOS Coagulation* study (n=284) (Table S2, Tab_1 for cohort descriptive data). In the *VEBIOS ER* and *Coagulation* study, mean CFHR5 concentrations in control plasma samples were 2842 ± 756 and 2467 ± 523ng/ml, respectively; levels in the range previously estimated by mass spectrometry (∼1900ng/ml) [37]. In the *VEBIOS ER* discovery study, the mean absolute CFHR5 concentration was confirmed as higher in patients with confirmed VTE, compared to patients where VTE was ruled out (3428 ng/ml ± 774 [cases] vs. 2842 ng/ml ± 756 [controls], p=1.05E-03 [age and sex adjusted]); the odds ratio (OR) for diagnosis of acute VTE associated with one standard deviation (SD) increase of CFHR5 concentration was 2.54 [confidence interval (CI) 1.52-4.66], p=1.05E-03 (Figure 1H.i). Consistent with the relative quantification results in our previous study [27], absolute CFHR5 concentration was associated with prior diagnosis of VTE in the extended *VEBIOS Coagulation* cohort, compared to controls (mean concentration 2681 ng/ml ± 554 [cases] vs. 2467 ng/ml ± 523 [controls], p= 8.42E-04 [age and sex adjusted]); the OR for first time VTE was 1.55 [CI 1.20-2.01], p=8.85E-04) (Figure 1J.i). We next investigated if CFHR5 levels were associated with VTE associated risk factors, such as age, body mass index [BMI]) and routine clinical laboratory tests for blood markers associated with thrombosis risk (e.g., D-dimer, c-reactive protein [CRP], thrombocyte count). In *VEBIOS ER*, CRP levels correlated with plasma CFHR5 concentration, in cases (ρ=0.57, p<0.001) and controls (ρ=0.52, p<0.001) (Figure 1H.ii), but there was no strong correlation between CFHR5 and the other parameters measured, in cases or controls, including D-dimer (Figure 1H.iii and Table S1, Tab_3, Table A). In the *VEBIOS Coagulation* study, CFHR5 levels in cases correlated with both CRP (ρ=0.49, p<0.001) and D-dimer (ρ=0.42, p<0.0001) (Figure 1J.ii and iii, right panel), but in controls these correlations were weak (CRP ρ=0.24, p=0.005) or absent, respectively (Figure 1J.ii, and iii, left panel and Table S1, Tab_3, Table B). The association between CFHR5 and VTE remained significant in both *VEBIOS ER* (p=0.0029) and *VEBIOS Coagulation* (p=0.0015) when adjusted for CRP. Therefore, we did not adjust for CRP in further analyses.

### CFHR5 is specifically expressed in liver hepatocytes with other complement related genes

To further understand the expression characteristics of *CFHR5*, and to identify possible co-expressed or co-regulated proteins we used a whole transcriptome analysis approach. In a consensus dataset, consisting of normalized mRNA transcript (nTPM) levels across 55 different human tissue types, generated from Human protein Atlas (HPA) (www.proteinatlas.org) [39] and Genotype-Tissue Expression Project (GTEx) (www.gtexportal.org) [40] datasets, *CFHR5* was highly and specifically expressed in the liver (Figure 2A). Single cell analysis of liver tissue [41], showed that *CFHR5* was specifically expressed in the hepatocyte cellular compartment (Figure 2B). To identify transcripts potentially co-expressed or co-regulated with *CFHR5* in the liver, we analysed bulk RNAseq data (n=226) retrieved from GTEx portal V8. We generated Pearson correlation coefficients between *CFHR5* and all other expressed protein coding transcripts (∼19,525) (Table S1, Tab_4, Table A) and used gene ontology (GO) and reactome analysis [42, 43] to identify over-represented classes and pathways in the top 50 most highly correlated genes (Table S1, Tab_4, Table B and C). Results were consistent with known CFHR5 function; significant GO terms included *‘complement activation*’ (False Discovery Rate, FDR =2.4E-16) and ‘*humoral immune response*’ (FDR =1.2E-12) and reactome pathways included ‘*regulation of complement cascade*’ (FDR=3.8E-24). We performed an unbiased weighted network correlation analysis (WGCNA) [44] on the same dataset, where correlation coefficients between all transcripts, excluding those classified as non-coding, were calculated and subsequently clustered into related groups, based on expression similarity (Table S1, Tab_4, Table A [column F]). Transcripts that fulfilled the criteria of: (i) Pearson correlation with *CFHR5* >0.65 and (ii) annotation to the *CFHR5*-containing gene cluster in WGNCA (group 68), were identified as those most likely to be co-expressed or co-regulated. Of these, 13/18 [72%] were other members of the complement cascade and 15/18 [83%] were also specifically expressed in liver [39] (Table S1, Tab_4, Table A, column F) (Table S1, Tab_4, Table A, column B, bold text). When these proteins were interrogated using the protein-protein interaction database STRING, v11 [45], 13/18 had high confidence functional and physical associations (Figure 2C) (Table S1, Tab_5, Table A). CFHR5 was most strongly linked to complement 3 (C3), the central hub of the largest of the three linked cluster groups identified (Figure 2C, clusters represented by green, red and cyan) (Table S1, Tab_5, Table B).

**Figure 2:**
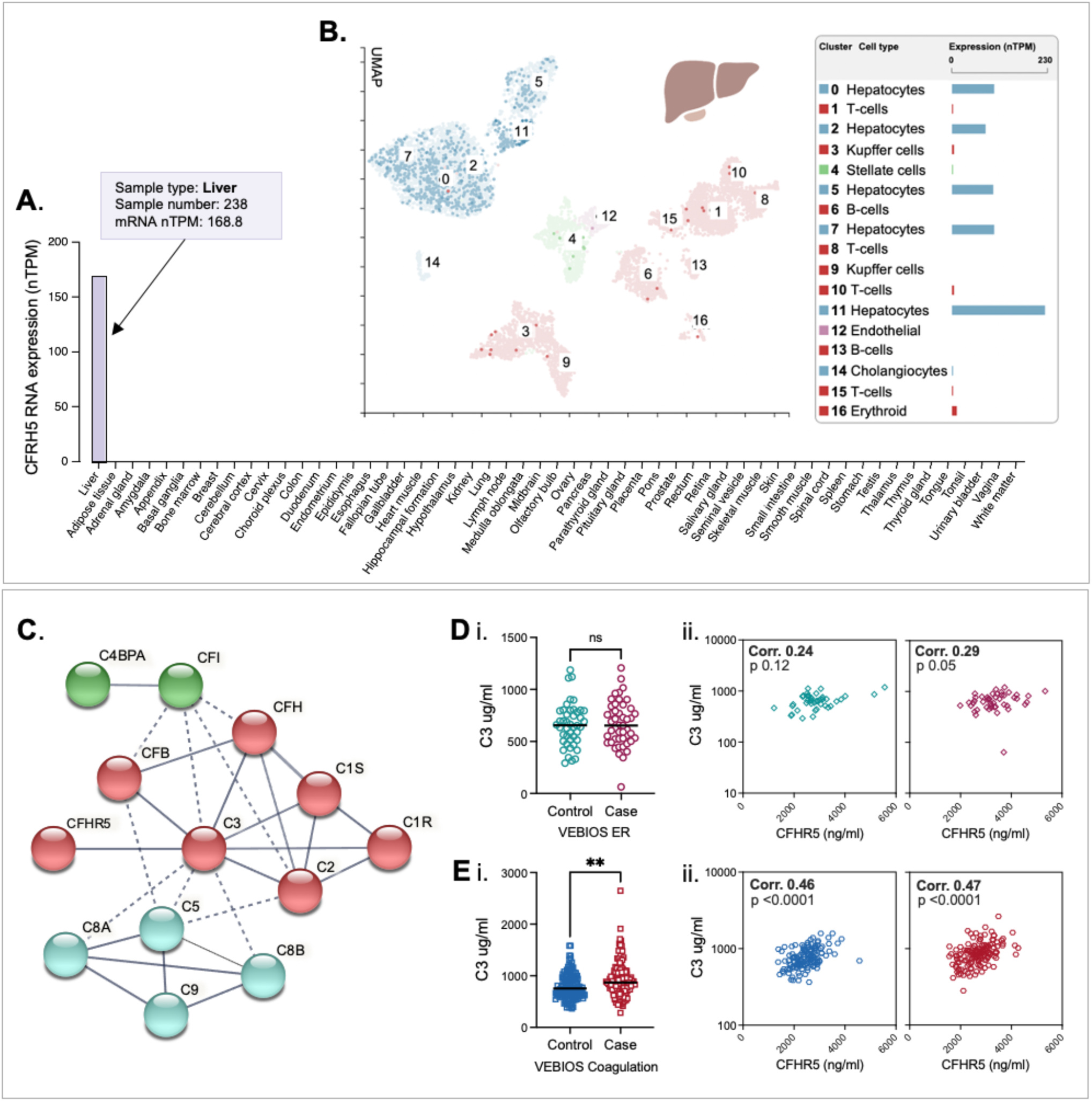
*CFHR5* is expressed in hepatocytes and is VTE-associated independent of C3. **(A)** mRNA expression of *CFHR5* across 55 different human tissue types. **(B)** Expression of *CFHR5* in different liver cell types, analysed by ssRNAseq. **(C)** STRING protein-protein interaction analysis for genes identified as potentially co-expressed with *CFHR5* in liver by correlation-based analysis of bulk mRNAseq. Coloured circles indicate closest network clusters. Complement component 3 (C3) concentration was measured in plasma from (**D)** *VEBIOS ER* or (**E**) *VEBIOS coagulation* to determine: (**i**) differences between controls and cases, or (**ii**) correlation with CFHR5 in controls (left) or cases (right). **p<0.01. C4BPA: complement factor 4 binding protein, CFI: complement factor, CFB: complement factor B, CFH; complement factor H, C1S; complement component 1, C1R; complement component 1, C2; complement component 2, C8a; complement component C8 alpha chain, C8b; complement component C8 beta chain, C5 complement component 5, C9; complement component 9

### CFHR5 is associated with VTE independent of C3

Plasma levels of C3 have previously been reported as associated with incident VTE [46]. To determine if the association between CFHR5 and VTE we observed is dependent or independent of the relationship with C3, we developed an in-house dual binder quantitative assay to measure C3 in the *VEBIOS ER* and *VEBIOS Coagulation* cohorts. In *VEBIOS ER*, plasma C3 was not elevated in cases, compared to controls (Figure 2D.i), CFHR5 and C3 did not significantly correlate in either group (Figure 2D.ii) and C3 was not associated with VTE (OR 1.04 [CI 0.68-158], p=0.86). Conversely, in *VEBIOS Coagulation* C3 levels were higher in plasma from cases, compared to controls (Figure 1E.i), and CFHR5 and C3 correlated with each other in both ([controls ρ= 0.46 p=<0.0001], [cases ρ = 0.47 p=<0.0001]). After adjusting for age and sex, one SD increase in C3 level was significantly associated with VTE (OR 1.53 [CI 1.18-2.01], p=1.93E-03). When adjusting for C3 together with age and sex, one SD increase in CFHR5 level was still associated with VTE in both *VEBIOS ER* (OR 2.65 [CI 1.53-5.01], p=0.00126) and *VEBIOS Coagulation* (OR 1.37 [CI 1.03-1.83], p=0.032). In contrast, when adjusting for CFHR5 together with age and sex, C3 was no longer significantly associated with VTE in *VEBIOS Coagulation* (OR 1.31 [CI 0.99-1.78], p=0.0645). Thus, CFHR5 is associated with both a diagnosis of acute VTE and prior VTE, independent of C3.

### The CFHR5 association with VTE replicates in additional cohorts

The identification of biomarkers associated with VTE diagnosis, or risk profiling, requires replication in independent cohorts, from different settings with different demographic profiles, to determine feasibility for potential translation to clinical practice. We sourced three independent replication cohorts to test the association of CFHR5 with VTE; the Swedish Karolinska Age Adjusted D-Dimer study (*DFW-VTE*) VTE study (n=200) consisting of patients sampled at presentation with suspected VTE at Huddinge hospital, Stockholm that was either subsequently confirmed (cases; n=54) or ruled out (controls; n=146) [47] (Figure 3B), the French *FARIVE* case/control study (n=1158) consisting of patients sampled during the week following a diagnosis of acute VTE (n=582), with hospital-based controls (n=576) [48] (Figure 3C) and the Spanish *Riesgo de Enfermedad TROmboembólica VEnosa* (*RETROVE*) study (n=668) of patients sampled post anticoagulant treatment (n=308), with population based controls (n=360) [49] (Figure 3E) (for all cohort details see Table S2, Tabs 2-4). The relative risk of VTE associated with CFHR5 per 1 SD increase in CFHR5 concentration was significant in all 3 replication cohorts: *DFW-VTE* (OR 1.80 [CI 1.29-2.58], p=0.0008) (Figure 3B), *FARIVE* (OR 1.24 [CI 1.10-1.40], p=0.0004) (Figure C), *RETROVE* (OR 1.29 [CI 1.09-1.53], p=0.002) (Figure 3E) (Table 2). In all cohorts, including *VEBIOS ER* and *VEBIOS Coagulation*, association results were comparable in sub-set analysis by thrombosis type (Table S1, Tab_6, Table A) or sex (Table S1, Tab_6, Table B). When samples from cases and controls were stratified according to CFHR5 concentration, the association with VTE risk was most pronounced in the third tertile, in all 5 cohorts analysed (Table 2).

**Figure 3:**
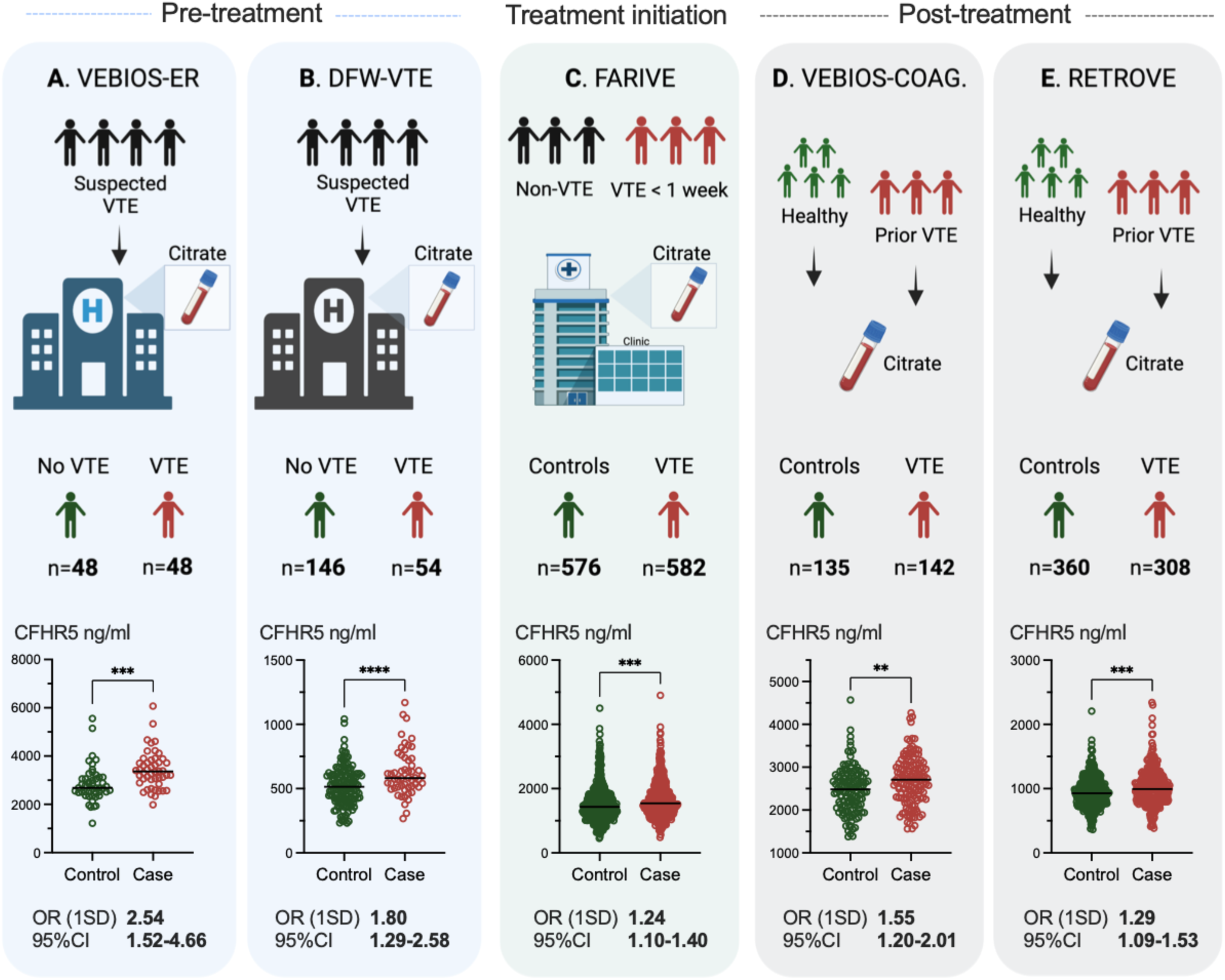
CFHR5 concentration is associated with VTE in 5 independent studies. Plasma samples were generated as part of: (**A**) the Swedish *VEBIOS ER* or (**B**) the Swedish *DFW-VTE* study, both of which recruited patients presenting with suspected VTE. Samples were drawn pre-treatment, and cases and controls were identified based on confirmed or ruled out diagnosis. (**C**) The French *FARIVE* study recruited patients with confirmed acute VTE, with controls recruited from hospital patients treated for non-VTE causes. Samples were drawn within one week from diagnosis, during initiation of treatment. (**D**) The Swedish *VEBIOS Coagulation* or (**E**), Spanish *RETROVE* study recruited cases from patients who had a prior first time VTE, sampled post-treatment (6-12 months anticoagulants), with healthy controls recruited from the general population. CFHR5 concentration was measured in the respective samples using a dual binder assay **p<0.01, ***p<0.001, ****p<0.001 OR (1SD) = Odds ratio for 1 standard deviation elevation. CI= confidence interval.

**Table 2.**
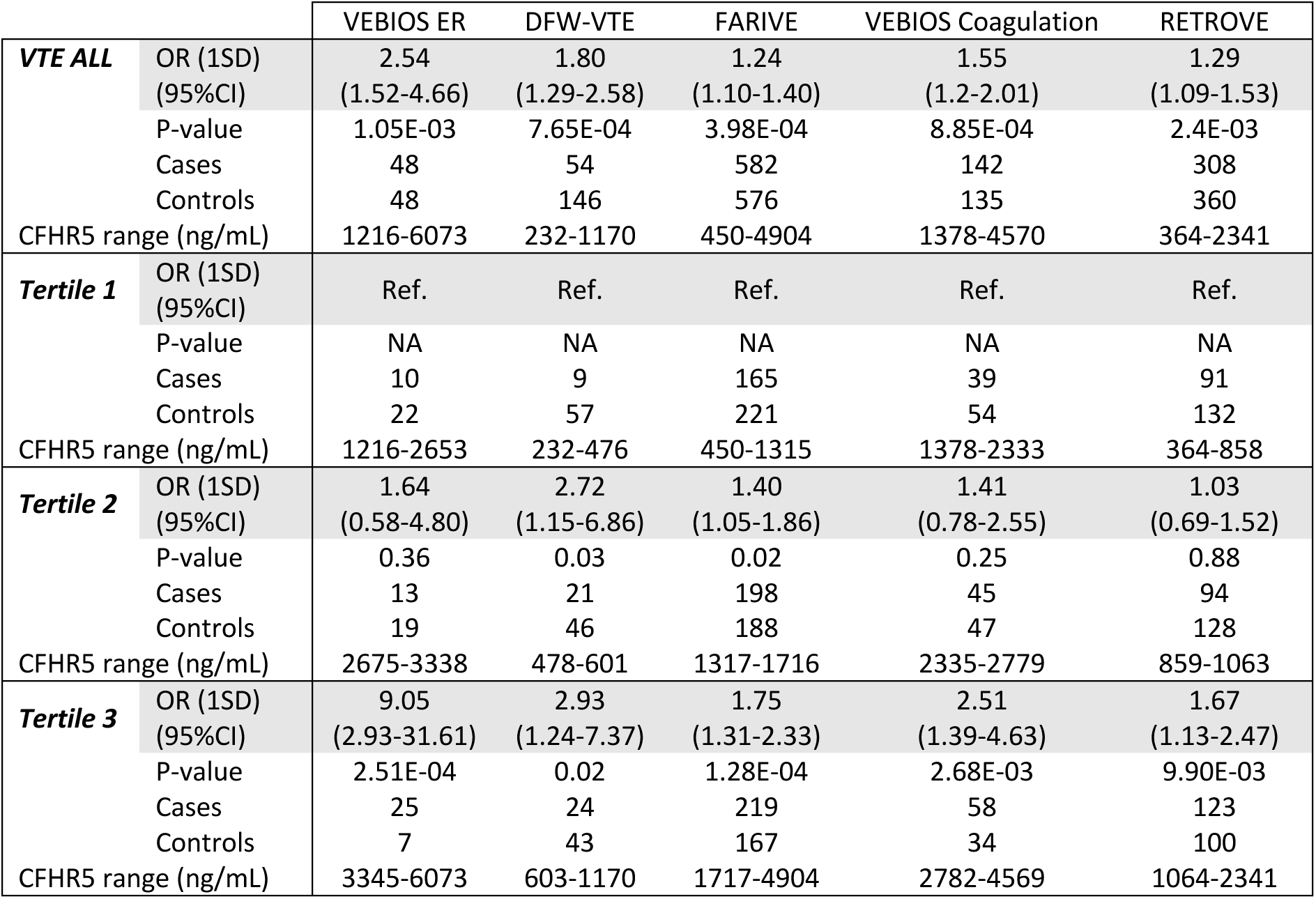
Risk of VTE associated with CFHR5 concentration in five independent studies.

### CFHR5 and risk of recurrent VTE

The MARseille THrombosis Association Study (MARTHA) is a hospital-b ased cohort of over 1,500 unrelated individuals recruited at the Thrombophilia center of La Timone hospital (Marseille, France). All patients had a history of a first VTE documented by venography, Doppler ultrasound, angiography and/or ventilation/perfusion lung scan [50]. We measured plasma CFHR5 concentrations in samples from VTE patients where follow up data was available (n=669) (for cohort details see Table S2, Tab_5). Using a Cox survival model with left truncature on the VTE cases we tested for association between CFHR5 and recurrence. In total, there were 124 recurrent events (52 among 231 males, 72 among 438 females). After adjusting for sex, familial history of VTE, provoked or unprovoked status of the first VTE, age at first VTE, and BMI, the Hazard Ratio (HR) associated of 1 SD increase in CFHR5 levels was HR = 1.13 [0.96-1.32], p = 0.134. The association was consistent between females (HR = 1.11) and males (HR = 1.14) and between patients with DVT (HR = 1.18) or PE as first event (HR=1.13). This association was driven by the subgroup of patients with unprovoked first VTE (HR = 1.32 [0.99 – 1.77], p=0.056), as no association was observed when the first event was provoked (HR = 1.01 [0.83 – 1.23], p=0.900). However, the testhomogeneity between these two HRs did not reach 0.05 significance (p = 0.23).

### Genome Wide Association Study on CFHR5 plasma levels

To explore if CFHR5 concentration in plasma was influenced by genetic variants, we performed a meta-analysis of GWAS data CFHR5 concentrations in individuals from the *FARIVE* (n=1,033), *RETROVE* (n=668) and *MARTHA* (n=1,266) studies. The results from the association results are summarised in Figure 4A. Of N= 7,135,343 SNPs tested in a total sample of 2967 individuals, one genome-wide significant (p < 5E-08) signal was observed on chr1q31.3. The lead SNP at this locus was rs10737681, mapping to CFHR1/CFHR4, and the G allele was associated with β= +81.33 ± 8.67 (p = 6.49E-21) increase in CFHR5 levels. After conditioning on the rs10737681, a borderline significant association (p = 9.83E-08) was observed at the rs10494747 where the A-allele tended to associate with an increase of β= +47.60 ± 8.92 in CFHR5 levels. Rs10494747 is distant of ∼343 kb from CFHR2 rs10737681, maps to ZBTB41 (Figure 4B) and was in moderate linkage disequilibrium (r^2^∼0.10, D’ = - 0.63) with the lead rs10494747 variant. Together, these two variants only explained 2.2% of the inter-individual variability in CFHR5 plasma levels. Neither SNP was associated with VTE risk in the *FARIVE* and *RETROVE* case-control samples (Table S1, Tab_7).

**Figure 4:**
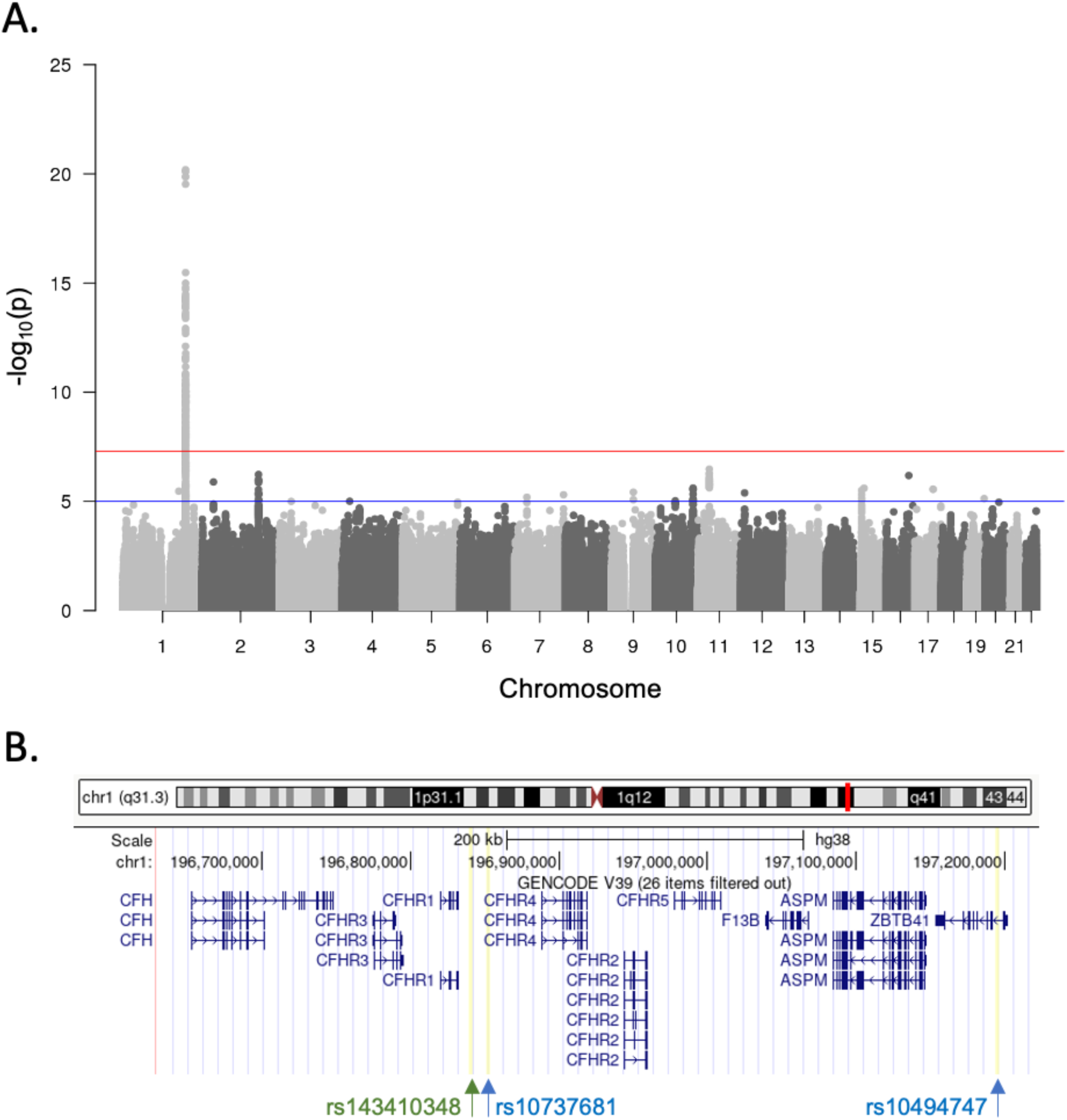
GWAS analysis identifies a CFHR5 pQTL on Chromosome 1 q31.3. A meta-analysis of GWAS data for 7,135,343 SNPs tested for association with CFHR5 concentrations in a total sample of 2967 individuals from the *FARIVE*, *MARTHA* and RETROVE studies: (**A**) identified one genome-wide significant (p<5E-08) signal on chr1q31.3. (**B**) The lead SNP at this locus, rs10737681, maps between the *CFHR1* and *CFHR4* gene loci in the gene cluster of *CFHR1*-*5*. A borderline significant association (p = 9.83E-08) with CFHR5 levels was observed at the rs10494747, mapping to the *ZBTB41* gene. Indicated in green is the rs143410348 recently identified with genome wide significance as associated with VTE risk [68].

### CFHR5 enhances platelet activation and degranulation in plasma

C3a, generated by cleavage of C3, can increase platelet activation [51–53]. As CFHR5 has a regulatory role upstream of C3/C3a activation, we investigated the effect of recombinant CFHR5 on platelet activation *in vitro.* Platelet rich plasma was pre-incubated with 6µg/ml recombinant CFHR5, a concentration corresponding to the upper range of that detected in the plasma of the VTE case group in *VEBIOS ER*, and in agreement with what has been reported in previous literature [54]. Platelet activation was measured by surface expression of P-selectin, activated GP IIb/IIIa or CD63 (Figure 5A, B and C, respectively) in response to adenosine diphosphate (ADP), convulxin or TRAP6 (Figure 5 i, ii and iii, respectively). Under baseline conditions (in the absence of stimulation) activation was not modified by pre-incubation with CFHR5 (Figure 5A.i-iii). Following stimulation with ADP, a higher percentage of platelets pre-incubated with CFHR5 expressed P-selectin (Figure 5A.i) (p=0.0056), activated GP IIb/IIIa (Figure 5B.i) (p=0.031) and CD63 (Figure 5C.i) (p=0.009), compared to the control. Pre-incubation with CHFR5 also potentiated platelet activation in response to convulxin or TRAP6 stimulation (Figure 5A-C.ii and iii), although the effect appeared to be more strongly linked to stimulus concentration, than that observed for ADP. Washed platelet response to ADP, convulxin or TRAP6 was not modified by preincubation with CFHR5 (Figure S2) (ANOVA all p>0.05), indicating that additional components in plasma are required for the observed effects, and that they are not a direct effect of CFHR5 on platelets. These data are consistent with the concept that CFHR5 is involved in the underlying pathology of VTE.

**Figure 5:**
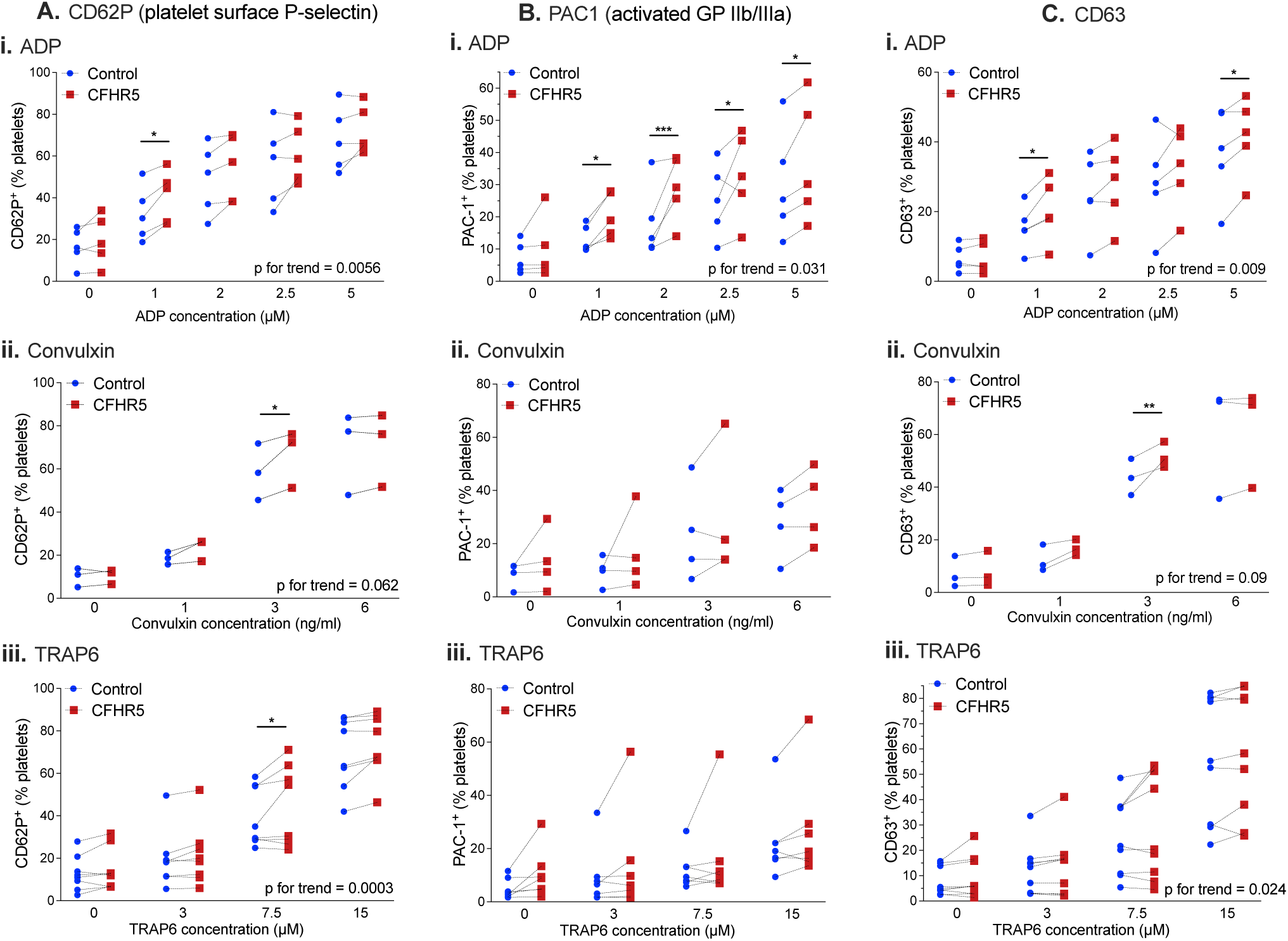
Recombinant CFHR5 enhances platelet activation in platelet rich plasma. Platelet activation was measured by surface expression of (**A**) P-selectin, (**B**) activated GP IIb/IIIa or (**C**) CD63, following treatment of platelet rich plasma with different concentrations of (**i**) adenosine diphosphate (ADP) (**ii**) convulxin or (**iii**) TRAP6, following pre-incubation (10 minutes) with recombinant CFHR5, or PBS control. Each experiment is represented by an individual point and paired experiments connected by a dotted line. *p<0.05 **p<0.01 ***p<0.001 (p for trend bottom right).

### CFHR5 is associated with thrombin generation potential in patients with previous VTE

As thrombin generation has been associated with increased risk of VTE [55], we tested the association between CFHR5 plasma concentration and thrombin generation as measured by thrombinoscope in *MARTHA* (n=774 VTE cases, see Table S2, Tab_5 for details) with replication in *RETROVE* (308 cases/360 controls, see Table S2_Tab_4 for details). In both *MARTHA* and *RETROVE cases*, we find significant association between CFHR5 and lag time (rho=0.181, p= p<0.0001 and rho=0.176, p<0.0001, respectively), Endogenous Thrombin Potential (ETP) (rho=0.105, p= 0.0036, and rho=0.130, p<0.0001, respectively), peak (rho=0.117, p= 0.0012, and rho=0.132, p<0.0001), and ttPeak (rho=0.116, p=0.0012, and rho=0.086, p=0.0274 (see Table S1, Tab_8).

### Plasma CFHR5 is associated with prognosis and respiratory status in COVID-19 patients

Accumulating evidence shows that vascular dysfunction, a prothrombotic state and complement activation underlies severe COVID-19 pathophysiology, manifested as respiratory failure linked to microvascular thrombosis in lung [56, 57]. Furthermore, VTE is a frequent complication in hospitalized COVID-19 patients [58]. To determine if CFHR5 was associated with COVID-19 severity, we measured relative plasma levels in 339 samples collected at consecutive timepoints from 112 hospitalised COVID-19 patients in the *COVID-19 biomarker and Immunity* (*COMMUNITY*) study [59] (Figure 6A) (cohort details see Table S2, Tab_6), using a single binder assay (antibody HPA073894). At each sampling timepoint, we categorised patients into one of four ‘*Respiratory support Index*’ (RI) groups, as a proxy variable for COVID-19 severity; **RI 0**: no respiratory support required, **RI 1**: ≤ 5 Litres of oxygen on nasal cannula or mask, **RI 2**: > 5 Litres of oxygen on nasal cannula or mask, **RI 3**: non-invasive ventilation/high-flow nasal cannula, or **RI 4**: intubation (Figure 6B). CFHR5 levels were positively associated with patient RI at the time of sample collection (p=<0.0001, ANOVA) (Figure 6C.i). To determine if the measurement of plasma CFHR5 at admission had potential value for prognosis, we performed a longitudinal analysis to determine the association with maximum respiratory support index (max RI) registered at *any* sampling time point during the hospital stay. CFHR5 at baseline was associated with the maximum level of respiratory support required (Figure 6C.ii) (p=<0.0022, ANOVA). Therefore, CFHR5 levels measured early upon admission carry short-term prognostic information of disease development.

**Figure 6.**
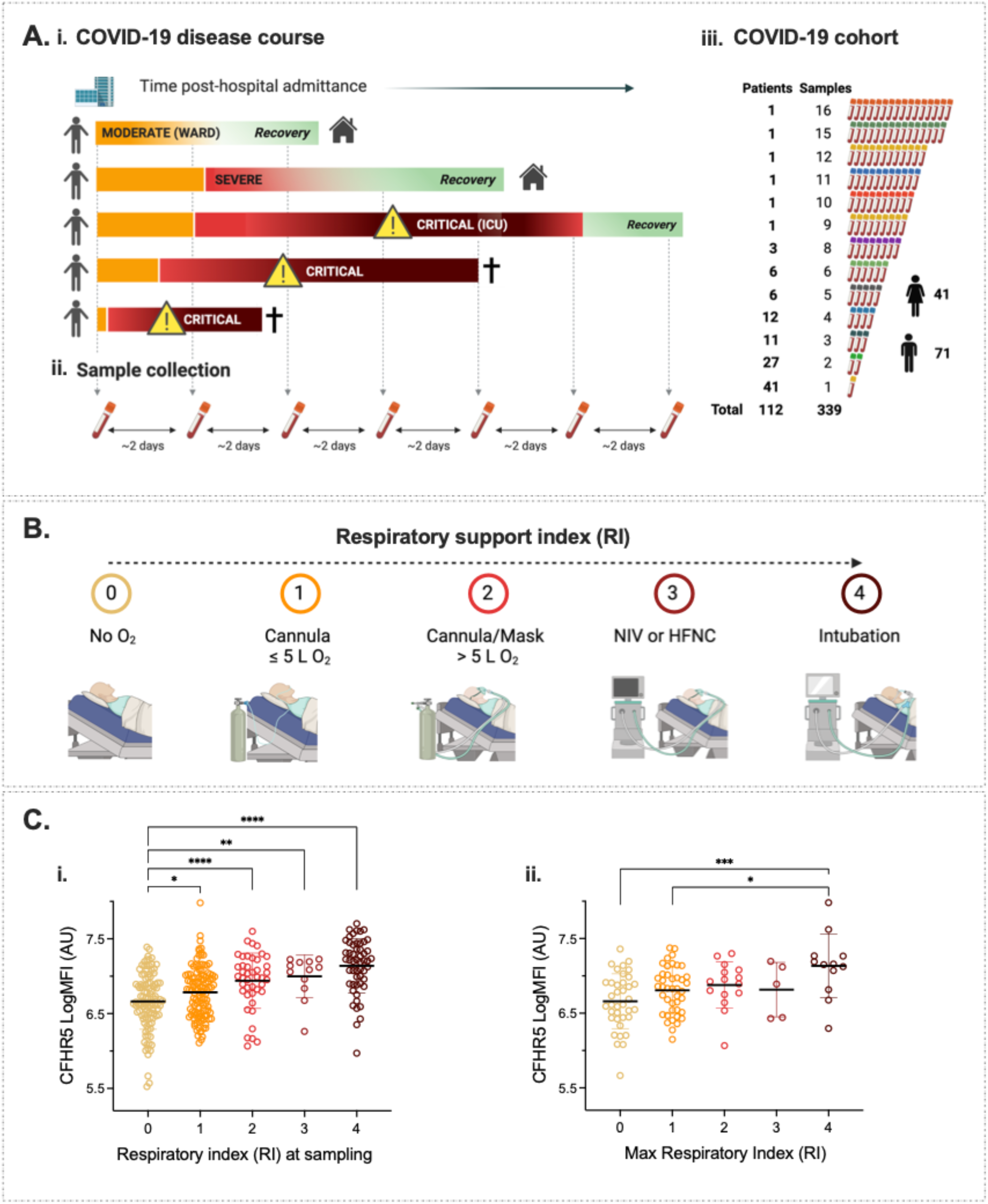
Plasma CFHR5 concentration is associated with COVID-19 severity. (**A**) Schematic representation of the *COMMUNITY* cohort; (**i**) patients hospitalised with acute COVID-19 were included prospectively and followed longitudinally for disease severity (**ii**). Plasma was sampled at inclusion and, on average, every second day during hospital stay. (**iii**) In total, 359 samples were collected from 112 patients, ranging from 1-16 samples per patient. (**B**) Disease severity registered at time of sampling was classified into a Respiratory support Index (RI): RI=0: no Oxygen (O_2_), RI=1: ≤ 5 litres (L) of O_2_ administered via cannula, RI=2: ≥ 5 L O_2_ via cannula or face mask, RI=3: Oxygenated air via Non-Invasive Ventilation (NIV) or High Flow Nasal Cannula (HFNC), RI=4: Intubated on ventilator. (**C**). CFHR5 plasma concentrations were measured in (i) all 339 samples to analyse the relationship with respiratory support index (RI) recorded at the time of sampling or (**ii**) in the 112 baseline samples to analyse the relationship with the maximum RI for that patient recorded at any subsequent time point (**ii**). *p<0.05, **p<0.01, ***p<0.001, ****p<0.0001.

## DISCUSSION

Here, we aimed to identify biomarkers associated with acute VTE that are linked to disease pathogenesis. Using a nested case-control study, derived from a cohort of patients presenting to the ER with suspected acute VTE, and from a case-control study with patients that had suffered a previous first VTE, we identify CFHR5, a regulator of the alternative complement activation pathway, as such a biomarker. The association of CFHR5 with current or previous VTE was replicated in three additional cohorts or case-control studies, and we also found a trend for association with risk for recurrence of unprovoked VTE. We identify a genome-wide significant signal associated with CFHR5 levels located in the CFHR1-5 gene cluster loci. We further provide evidence of a direct role of CFHR5 in the induction of a pro-thrombotic phenotype, through its effect on platelet activation. Finally, we show that increased CFHR5 was associated with degree of respiratory insufficiency and prognosis in acute COVID-19. Our findings indicate CFHR5 has potential application as a clinical biomarker for VTE diagnosis and risk prediction, providing further support to the idea that complement regulation is a key element of VTE pathogenesis.

Currently, D-dimer is the only plasma biomarker used in VTE diagnostic work-up, but its clinical utility is limited to ruling out VTE in low-risk patients. Several studies have attempted to identify novel biomarkers with potential clinically usefulness for the confirmation of VTE diagnosis, and although a number have been identified [11], none have yet been implemented in clinical practice. For many, like D-dimer, elevated levels are a consequence of thrombosis formation, e.g. biomarkers of fibrinolysis, clot re-modelling or resolution (e.g. MMPs), inflammation secondary to local vascular and tissue injury (e.g., CRP, IL-6, IL-10, fibrinogen), or of endothelial and/or platelet activation (e.g. vWF, P-selectin) [11, 60, 61].

We find no correlation or association between D-dimer and CFHR5 in the acute VTE setting, supportive that increased concentration at diagnosis is not secondary to thrombus formation. In contrast, we found a strong correlation between D-dimer and CFHR5 levels in patients followed up after ending treatment for a first VTE, but not in controls. D-dimer has been associated with increased risk of first and recurrent VTE [12–18, 62] and thus, our results are consistent with a link between CFHR5 and subclinical coagulability in these patients at follow-up, possibly due to persistent risk factors.

CFHR5 shares sequence and structural homology with Complement Factor H (CFH) [63], the main negative regulator of alternative pathway (AP) activation in plasma [64]. Under normal conditions, the AP is constitutively active through spontaneous hydrolysis of the thioester bond in C3 and the formation of the initial fluid phase C3 convertase, C3(H_2_O)Bb, which cleaves C3 into C3a and C3b [64]. CFH promotes decay of the alternative and classical pathway convertases and is a cofactor in the cleavage of C3b, hereby regulating excess activation of C3 [65]. CFH inhibits C3 convertase by competing with Complement Factor B (CFB) for binding to C3, preventing formation of C3a. CFHR5 antagonizes CFH function, through competitive binding to C3b and its fragment C3d [66], thus deregulating AP activation. CFHR5 also promotes complement activation by interfering with CFH binding to CRP, pentraxin 3 (PTX3), and extracellular matrix (ECM) [67]. The complement and haemostatic systems interact at several points during initiation, propagation, and regulation of complement activation and coagulation [68]. Studies have indicated a role of complement in VTE pathogenesis [46, 52], but underlying mechanisms are not well understood. Elevated plasma C3 in baseline samples has been shown to be associated with increased risk of future VTE [46]. Consistent with these findings, C3 was associated with prior VTE in the *VEBIOS coagulation* study, but not with acute VTE in the *VEBIOS ER* study. In both cases, and in the previous study by Nordgaard *et. al.* [46], total C3 level, rather than the active form (C3a) is measured; it is possible that in acute VTE, regulation of C3 convertase (by CFHR5) is important, rather than absolute C3. It could be speculated that the association of C3 with VTE in individuals sampled pre-VTE [46] or following treatment, reflects co-regulation of CFHR5 and C3 under basal conditions, where C3 is a proxy for CFHR5, where CFHR5 represents the functional link to VTE risk, rather than C3 levels *per se*. Consistent with this idea, in the VEBIOS coagulation study, the association between CFHR5 and previous VTE remained significant when adjusted for C3.

The *CFHR5* locus maps to chromosome 1q31.3 at one end a gene cluster that spans approximately 350 kb and that includes (in order from *CFHR5*) the *CFHR2, CFHR4, CFHR1, CFHR3*, and *CFH* loci (Figure 4B). The rs10737681 we identify with genome wide association with CFHR5 levels maps between the CFHR4 and CFHR1 genes (Figure 4B). Of note, the *CFHR2* locus has just been identified as a novel susceptibility locus for VTE in the recently published international effort on VTE genetics [21]. The lead SNP at this locus is rs143410348, which is in moderate LD with the rs10737681 (r^2^ = 0.38 in *FARIVE*) that here we found associated with CFHR5 plasma levels. Unfortunately, rs143410348 was not imputed in *MARTHA* nor in *RETROVE*, preventing us from investigating its association with CFHR5 plasma levels in our meta-analysis. Nevertheless, in *FARIVE* where it was imputed, its association with CFHR5 levels tended to be slightly less pronounced than that observed with rs10737681 (+76.4 ± 27.0; p = 4.8E-03 vs +101.5 ± 29.1; p = 5.06E-04). Altogether, these observations emphasize the need for a deeper investigation of the genetic architecture of the *CFHR1*/*CFHR4*/*CFHR2*/*CFHR5* locus with respect to VTE risk.

Our study indicates that CFHR5 has a possible role in the potentiation of platelet activation. Using a flow restricted tissue-factor dependent mouse model of VTE, Subramaniam *et. al.* showed that C3 and C5 affected platelet activation and tissue factor procoagulant activity by different mechanisms, independent of formation of the terminal complement C5b-C9 complex [53]. In ligated inferior vena cava, both C3 deficient and C5 deficient mice developed smaller thrombi, with reduced fibrin deposition compared to the wild type. C3, but not C5, deficient mice had reduced platelet activation *ex vivo*, reduced platelet deposition *in vivo*, and reduced thrombosis incidence (<30 vs. 80% in wild type). These data indicate that in VTE C3 has an important role in initial hemostasis, independent of downstream complement proteins [52]. C3a, acting through platelet receptor C3aR, has an important role in the activation of the Gp3a/2b fibrinogen receptor via intraplatelet signaling, and subsequent thrombosis formation [51]. Similar to the study of Subraminam *et. al.* [53], Sauter *et. al*. showed C3 deficient mice had prolonged bleeding time, that could be reversed by intravenous administration of C3a peptide [51]. The C3a-C3aR induced intracellular signaling was mediated through the Rap1b activation, where co-stimulation of platelets with C3a-ADP resulted in increased Rap1b activity on top of the platelet stimulation by only ADP. In our study, we observe a similar co-stimulatory effect of CFHR5 on ADP- (and convulxin- or TRAP6-) induced platelet activation. This effect was observed on platelets in plasma, but not on those that were pre-washed, consistent with the effect of CFHR5 on platelet activation being due to its interaction with other complement factors (i.e., C3) in plasma. On basis of these recently published mechanistic findings, our *in vitro* results indicate that CFHR5 regulation of the alternative pathway of activation has an important role in C3a mediated platelet activation in thrombosis, providing a potential functional link to its association with acute VTE. Our findings indicate that CFHR5 mediated dysregulation of the AP could play an important role in COVID-19 progression. Acute respiratory distress syndrome (ARDS) is the most common serious complication of COVID-19 [69], where patients suffer from dysregulated haemostasis, presenting with atypical ARDS [69, 70] with thrombosis formation in the lung microvasculature [71]. Other studies have shown that severe disease is associated with complement activation [56], and the cross-talk between coagulation and complement contributes to elevated coagulopathy and thromboembolic complications [72]. Furthermore, elevated complement activation has been correlated with disease severity and development of ARDS in hospitalised COVID-19 patients [72] and patients with severe COVID-19 have a higher incidence of VTE [73].

Our study has various strength and limitations; *VEBIOS ER*, the discovery cohort, that was derived from a single centre, where blood sampling for plasma biobanking was performed in parallel to that for routine tests after initial evaluation (before diagnostic imaging or anticoagulant treatment), thus avoiding bias in inclusion or biobanking. Samples were handled according to standard clinical chemistry lab routine, thus variations in needle-to-spin-to-freeze time were equivalent between case and control samples. As biobanking was based on the routine sample flow, this increases the feasibility that identified biomarker candidates are suitable for clinical translation into a routine setting. Importantly, we demonstrate an association of CFHR5 with VTE in several independent studies, that include patients in the acute setting, at follow up, and prior to recurrence. One limitation of our study is that we have not analysed a cohort of individuals that were sampled prior to VTE event. From a technological perspective, our study demonstrates the need for orthogonal verification of any potential biomarker identified using antibody-based proteomics screening [34, 35]. The same caution should be extended to findings generated using other high throughput affinity proteomics technologies vulnerable to non-specific protein binding, such as aptamer-based [74], where missense single nucleotide polymorphisms can affect binding in a manner where a genetic difference drive associations, rather than protein levels (Figure S1 F) [75, 76]. Studies comparing different affinity proteomics technologies have found correlations of proteins assayed with two or more platforms to range from highly concordant (r = 0.95) to inversely correlated (r = −0.48) [75], highlighting further the need for orthogonal validation of any potential biomarker identified.

The next steps towards the translation of our findings into a clinical setting is to develop and establish standardised methods for quantification, to establish reference intervals and define cut off values with respect to specificity and sensitivity. Furthermore, it remains to be established if the incorporation of CFHR5 measurements into clinical decision rules or other scores can improve predictive power. The inclusion of CFHR5 measurements as a diagnostic and/or risk predictive marker in randomized clinical trials of acute VTE and VTE recurrence would be particularly informative, as these are two areas of high clinical relevance in need of improved tools for clinical decision making. Furthermore, while our study indicates that CFHR5 has a functional role in VTE development, further studies are needed to understand the mechanism.

## MATERIALS AND METHODS

### PATIENTS AND SAMPLES

#### Discovery study

##### Venous thromboembolism biomarker study (VEBIOS)

*VEBIOS* is part of a collaboration between Karolinska University Hospital, Karolinska Institute and Royal Institute of Technology (KTH) designed to identify new plasma biomarkers for VTE [27]. *VEBIOS* comprises two different studies: (**i**) *VEBIOS ER study* is a prospective cohort study carried out at the Emergency Room (ER) at the Karolinska University Hospital in Solna, Sweden, between December 2010 and September 2013. All patients admitted with the suspicion of deep vein thrombosis (DVT) in the lower limbs and/or pulmonary embolism (PE), over 18 years old were eligible for the study. Exclusion criteria were patients with on-going anticoagulant treatment, pregnancy, active cancer, short life expectancy or lack of capacity to leave approved consent. A case was defined if a) VTE was confirmed by diagnostic imaging - compression venous ultrasonography (CUS) in patients with suspected DVT in the lower limbs, and computed tomography pulmonary angiography (CTPA) in patients with suspected PE, and b) anticoagulant treatment was initiated based on the VTE diagnosis. Patients with no evidence of an acute VTE, (neither by diagnostic imaging nor by Well’s clinical criteria) that had a normal d-dimer test [5], were referred as controls in the study. All participants were sampled before any anticoagulant treatment. Whole blood was collected in citrate or EDTA anticoagulant at the ER and sent within 30 minutes to the Karolinska University Laboratory. After centrifugation at 2000*g* for 15 minutes, plasma aliquots were snap frozen and stored at -80°C. *Data collection*: For each patient, doctors filled in a questionnaire detailing (1) any provoking factors within one month preceding the visit to the ER (2) current health situation, alcohol consumption and smoking habits; (3) family history of VTE (4) ongoing antithrombotic (antiplatelet) treatment and (5) estrogen containing contraceptives and hormone replacement therapy (women only). Information from the ER visit on patient weight and height (when available) along with results from routine laboratory tests i.e., blood count, d-dimer, C-reactive protein (CRP), creatinine, international normalized ratio (INR) and activated partial thromboplastin time (aPTT) were extracted from the medical records. In total, 158 patients were included (52 cases). For the present study, 48 cases were available for analysis and 48 controls were matched as closely as possible by age and sex. Clinical characteristics of the sample set is given in Table 1. (**ii**) *VEBIOS Coagulation study* is an on-going case-control study established January 2011 of patients sampled at an outpatient coagulation clinic after discontinuation of anticoagulant treatment after a verified first VTE (DVT to the lower limbs and/or PE), matched with healthy controls from the population. Patients were between 18 to 70 years of age, free from cancer, severe thrombophilia and pregnancy at inclusion [27]. In the present study, we analysed an extended sample set of *VEBIOS Coagulation* comprising all available samples; 144 cases and 140 controls (Table S2, Tab_1). Approval for *VEBIOS* was granted by the regional research ethics committee in Stockholm, Sweden (KI 2010/636-31/4) and all participants gave informed written consent, in accordance with the Declaration of Helsinki.

#### Replication cohorts

The Swedish Karolinska Age Adjusted D-Dimer study (*DFW-VTE study*) includes patients with clinically suspected acute VTE, prospectively recruited from the ER of Karolinska University Hospital in Huddinge, Stockholm, as previously described [47]. The patients were out-patients with low-to-high probability of acute PE or DVT in a lower limb. The study was approved by the regional ethics review board in Stockholm (DNR 2013-2143-31-2), and all participants gave informed written consent, in accordance with the Declaration of Helsinki. For the current study, biobanked plasma aliquots were available for a subset of subjects comprising 15 patients with PE, 39 with DVT, and 146 controls where VTE was excluded. Controls were identified based on negative diagnostic imaging, or a low Welĺs score together with negative D-dimer. Clinical characteristics are described in Table S2, Tab_2.

The *FARIVE study* is a French multicentre case-control study carried out between 2003-2009, as previously described [77]. The study consists of patients with first confirmed VTE (DVT to the lower limbs and/or PE) from 18 years of age, matched to hospital controls with no previous thrombotic event. All participants were free from cancer. The study was approved by the Paris Broussais-HEGP ethics committee in Paris (2002-034) and all participants gave informed written consent, in accordance with the Declaration of Helsinki. In the current study we used a subset of *FARIVE* samples (n=1158), as previously described [27] [78]. From most cases, blood was collected in the first week after diagnosis and during anticoagulant treatment initiation. Clinical characteristics are described in Table S2, Tab_3.

The *Riesgo de Enfermedad TROmboembólica VEnosa (RETROVE) study* is a prospective case–control study of 400 consecutive patients with VTE (cancer associated thrombosis excluded) and 400 healthy control volunteers. Individuals were recruited at the Hospital de la Santa Creu i Sant Pau of Barcelona (Spain) between 2012 and 2016. Controls were selected according to the age and sex distribution of the Spanish population (2001 census) [79]. All individuals were ≥ 18 years. All procedures were approved by the Institutional Review Board of the Hospital de la Santa Creu i Sant Pau, and all participants gave informed written consent, in accordance with the Declaration of Helsinki. In the current study, samples from 308 cases and 360 controls were used. Clinical characteristics are described in Table S2, Tab_4.

The *Marseille Thrombosis Association study* (*MARTHA*) is a population based single centre study, as previously described [78]. Recruitment in *MARTHA* started in 1994 at Timone Hospital in Marseille (France) and is still ongoing. The cohort from 1994 and 2008, includes a total of 1542 VTE-cases (66% women) that donated blood for further analysis. Ethical approval was granted from the Department of Health and Science, France (2008-880 & 09.576) and all participants gave informed written consent, in accordance with the Declaration of Helsinki. In the current study, proteomics data generated for 1322 sampled *MARTHA* cases was used. For 669 of the *MARTHA* cases, follow up data up to 12 years post-event was available and used to analyse risk of recurrent VTE. For a subset of 774 cases data for thrombin generation potential (TGP) was available [80], which was used to analyse the association between CFHR5 and blood coagulability. Clinical characteristics are described in Table S2, Tab_5.

The *COVID-19 biomarker and Immunity (COMMUNITY) study* is a single centre study of 112 patients with COVID-19 admitted to a general ward, intermediate units, or intensive care units at Danderyd Hospital, Stockholm Sweden between April 15^th^ and May 27^th^, 2020. Inclusion required confirmed SARS-CoV2 infection, based on viral RNA detection by reverse-transcriptase polymerase chain reaction of nasopharyngeal or oropharyngeal swabs, or clinical presentation of COVID-19. Exclusion criteria were age <18 years. Patients were followed longitudinally from inclusion, and blood samples were collected shortly after hospital admission and every 2-3 days during the hospital stay. Procedures for blood sampling and plasma preparation have been previously described [59]. Demographic data, routine lab results, comorbidity and information and variables, reflecting clinical deterioration, including respiratory support were obtained from medical records. Patients were divided into groups based on respiratory support required at the time of sampling (‘Respiratory support Index’ or RI): RI = 0; no respiratory support, RI = 1; ≤5L of oxygen on nasal cannula or mask, RI = 2; >5L of oxygen on nasal cannula or mask, RI = 3; non-invasive respiratory support and RI = 4; intubation. Level of respiratory support and oxygen supplementation were set at by the treating physician. For the current study, 339 samples collected from 112 patients were analysed, with at least 2 samples for 71 patients (63.4%). The study was approved by the National Ethical Board (EPM 2020-01653). Clinical characteristics are described in Table S2, Tab_6.

### ANALYSIS OF PLASMA BY TARGETED PROTEOMICS

Plasma proteomic profiles in *VEBIOS ER* were generated using multiplexed suspension bead arrays (SBA) with 758 antibodies selected from the Human Protein Atlas (HPA) project, targeting 408 proteins (Table S1), using identical design, procedures and methods as previously described [27]. Briefly, paired samples were randomly distributed within the same 96-well area. Two suspension bead arrays composed of 380 antibodies and 4 controls were used to sequentially generate profiles of the 96 samples in parallel. Proteomics profiling was performed in both EDTA and Citrate plasma. Median fluorescence intensity (MFI) values were obtained from the suspension bead array assay by detecting at least 32 beads per ID and sample with the FlexMap 3D instrument (Luminex^®^ Corp) and were normalized by (1) probabilistic quotient normalization as accounting for any potential sample dilution effects [81], and (2) multidimensional MA (M=log ratio; A=mean average, scales) normalization to minimize the difference amount the subgroups of the samples generated by experimental factor as multiple batches [82]. Association of target proteins with VTE was tested using linear regression analysis while adjusting for age and gender. Log-transformation was applied to reduce any skewness in the proteomic data distribution. The analyses were performed using the R statistical computing software [83].

#### Immunocapture mass spectrometry (IC-MS)

IC-MS was performed in triplicate, as previously described [27] using the HPA059937 antibody (Atlas Antibodies) or rabbit immunoglobulin G (rIgG) as a negative control. Digested samples were analyzed using an Ultimate 3000 RSLC nanosystem (Dionex) coupled to a Q-Exactive HF (Thermo). Uniprot complete human proteome (update 20180131) was used to query the raw data, with the engine Sequest and Proteome Discoverer platform (PD, v1.4.0.339, Thermo Scientific). An internal database containing the most common proteins detected by IC-MS in plasma was used to calculate Z-scores [36].

#### In-house developed bead based dual binder immunoassays

A Suspension Bead Array (SBA) was built with the capture antibodies raised against human extracellular sulfatase 1-SULF1 (rabbit polyclonal HPA059937) and human CFHR5 (rabbit polyclonal HPA072446 and HPA073894) covalently coupled to color-coded magnetic beads as previously described (Drobin et al., 2013; Neiman et al., 2013). Bead-coupled rabbit IgG (Bethyl Laboratories Inc.) and mouse-IgG and bare beads were included as negative controls. Mouse anti-human SULF1 antibodies Abnova ABIN525031, Abcam ab172404, Thermofisher PA5-113112, Human Protein Atlas antibodies HPA054728 and HPA051204, and mouse monoclonal anti-human CFHR5 (R&D systems, MAB3845) antibody were labelled with biotin and used as detection antibodies in combination with their respective capture antibodiesCitrate plasma samples were thawed on ice and centrifuged for 1 min at 2000 rpm and diluted in buffer polyvinyl casein 10% rIgG (PVXcas 10% rIgG; polyvinyl alcohol, Sigma Aldrich P8136; polyvinylpyrrolidone, Sigma Aldrich PVP360; Blocker Casein, Thermo 37528), heated at 56°C for 30 min and incubated with the SBA overnight. The detection antibody was used at 1ug/mL for 90 min, and streptavidin-R-phycoerythrin (R-PE) conjugate (Life Technologies; SA10044) was used for the fluorescence read out in Luminex platform. The dual binder assays based on HPA059937, HPA072446 or HPA073894 as capture antibodies together with the monoclonal anti-human CFHR5 were used to measure samples in the *VEBIOS ER* cohort, and correlations between MIF values obtained with the 3 different dual binder assays and the original single binder assay with HPA059937 were determined.

#### Absolute quantification of CFHR5, C3 and D-dimer

For CFHR5 quantification, rabbit polyclonal anti-human CFHR5 HPA072446 (Atlas Antibodies) and mouse monoclonal anti-human CFHR5 (R&D systems; MAB3845) antibodies were used in a dual binder assay. Human recombinant CFHR5 (R&D systems; 3845-F5) spiked into chicken plasma (Sigma Aldrich, St. Louis, United States, P3266) was used as a standard. All samples were diluted 1:300 in PVXcas 10% rIgG. For C3 quantification, mouse anti-human C3 and mouse monoclonal anti-human C3 antibodies (Bsi0263, Bsi0190, respectively, Biosystems International) were used in a dual binder assay. Human recombinant C3 (Sigma Aldrich, C2910) spiked into C3 depleted serum (Merck, 234403) was used as a standard. All samples were diluted 1:5000 and analysed as described above. D-dimer was quantified by ELISA (D-Di 96 test, product #00947, Asserachrom, France) following the manufactureŕs instructions. In the *DFW-VTE* study, D-dimer values were analysed at the Karolinska University Hospital Laboratory in fresh samples sent for routine clinical chemistry analysis, as part of the work up in the ER.

#### Analysis of CFHR5 in COVID-19 patients in the *COMMUNITY* study

The HPA antibody HPA073894, targeting the CFHR5 protein, was included in a screening panel using multiplexed suspension bead arrays (SBA) following a similar protein profiling protocol as in the targeted plasma proteomics screening (see above). Association of CFHR5 plasma levels with Respiratory support Index (RI) at baseline was tested using linear regression models. RI groups were as follows: RI 0: no respiratory support required, RI 1: ≤ 5 Litres of oxygen on nasal cannula or mask, RI 2: > 5 Litres of oxygen on nasal cannula or mask, RI 3: non-invasive ventilation/high-flow nasal cannula, or RI 4: intubation. To handle multiple time point measurements, association of CFHR5 plasma levels with RI was further investigated using all available longitudinal measurements using a linear mixed effect model as implemented in the *nlme* R package. Analyses were adjusted for age, sex and body mass index.

#### CFHR5 mRNA expression across human organs

As part of the Human Protein Atlas (HPA, www.proteinatlas.org/), the average TPM value of all individual samples for each human tissue in both the HPA and Genotype-Tissue Expression Project (GTEx) transcriptomics datasets were used to estimate the respective gene expression levels. To be able to combine the datasets into consensus transcript expression levels, a pipeline was set up to normalize the data for all samples. In brief, all TPM values per sample were scaled to a sum of 1 million TPM (denoted pTPM) to compensate for the non-coding transcripts that had been previously removed. Next, all TPM values of all samples within each data source were normalized separately using Trimmed mean of M values to allow for between-sample comparisons. The resulting normalized transcript expression values (nTPM) were calculated for each gene in each sample. For further details see www.proteinatlas.org/about/assays+annotation <u>-normalization_rna</u>. Analysis of liver single cell transcriptomes and visualisation was performed as part of the HPA single-cell transcriptomics map [41] from data generated in [84].

#### CFHR5 mRNA co-expression analysis

Liver bulk RNAseq data analysed in this study was part of the Genotype-Tissue Expression (GTEx) Project (gtexportal.org) [85] (dbGaP Accession phs000424.v7.p2) (n=226). Pearson correlation coefficients were calculated between *CFHR5* expression values and those for all other mapped protein coding genes across the sample set. Weighted correlation network (WGCNA) analysis: The R package WGCNA was used to perform co-expression network analysis for gene clustering, on log2 expression values. The analysis was performed according to recommendations in the WGCNA manual. Genes with too many missing values were excluded using the goodSamplesGenes() function. The remaining genes were used to cluster the samples, and obvious outlier samples were excluded. Using these genes and samples a soft-thresholding power was selected and the networks were constructed using a minimum module size of 15 and merging threshold of 0.05. Eigengenes were calculates from the resulting clusters and eigengene dendrograms were constructed using the plotEigengeneNetworks() function.

#### Measurement of Thrombin Generation Potential

Thrombin generation potential (TGP) was measured in fresh frozen platelet poor plasma (PPP) using the Calibrated Automated Thrombogram (CAT®) method according to the manufacturer’s instructions. Analyses in MARTHA and RETROVE are as previously described [80, 86].

Output parameters recorded were lagtime (min), the time to the initial generation of thrombin after induction; Endogenous Thrombin Potential (ETP)(nmol/min), equal to the area under the Thrombogram curve; Peak (nmol/L), the maximum amount of thrombin produced after induction by 5pM tissue factor.

#### Effect of recombinant CFHR5 on platelet activation

Blood was drawn from healthy volunteers free from any anti-platelet therapy for at least 10 days and anticoagulated with sodium citrate. All donors signed informed consent, in accordance with approval of the Human Ethics Committee of the Medical University of Vienna (EK237/2004) and the Declaration of Helsinki. Whole blood was centrifuged (120 *g*, 20 minutes, room temperature) and platelet-rich plasma (PRP) harvested. To obtain isolated platelets, PRP was diluted with PBS and treated with PGI_2_ (100 ng/ml), centrifuged for 90 sec at 3000 x g and platelets were resuspended in PBS. This step was repeated twice. Platelet-rich plasma (PRP) or isolated platelets were incubated with recombinant CFHR5 in PBS (6 µg/ml, 3845-F5, R&D systems) or PBS alone for 10 minutes before treatment with varying concentrations of ADP (1-5 µM), TRAP-6 (3-15 µM) or convluxin (1-6 ng/ml) for 15 minutes. Platelets were subsequently incubated with primary antibodies: anti-human CD62P-AF647 (AK4), anti-human CD63-PE (H5C6) or anti-human CD41/CD61-FITC (PAC-1) (all Biolegend) for 20 minutes, washed (PBS then 500 *g* for 10 minutes), then fixed with 1 % paraformaldehyde and incubated with Alexa Fluor 647-streptavidin (Jackson Immuno Research, Ely, UK) for 20 minutes. Samples were analysed by flow cytometry (Cytoflex, Beckman Coulter GmbH, Krefeld, Germany) and data processed using Cytexpert (Beckman Coulter GmbH, Krefeld, Germany).

#### Genome wide genotyping methods

*RETROVE* samples were typed with the Illumina Infinium Global Screening Array (GSA) v2.0 array at the Spanish National Cancer Research Centre in the Human Genotyping lab, a member of CeGen. After genotyping, all monomorfic and unannotated variants were removed as well as polymorphisms with call rate <95% and those whose genotype distributions deviate from Hardy-Weinberg equilibrium at p < 0.000001. Remaining polymorphisms were then imputed using the TOPMed r2 reference panel using Eagle v2.4. *FARIVE* participants were genotyped using the Illumina Infinium Global Screening Array v3.0 (GSAv3.0) microarray at the Centre National de Recherche en Génomique Humaine (CNRGH). A control quality has been performed on individuals and genetic variants using Plink v1.9 and the R software v3.6.2 (Chang et al., 2015). Individuals with at least one of the following criteria were excluded: discordant sex information (N=20), relatedness individuals (N=9) identified by pairwise clustering of identity by state distances (IBS), genotyping call rate lower than 99% (N=5), heterozygosity rate higher/lower than the average rate +/- 3 standard deviation (N=34). These criteria led to a final sample composed of 1,266 individuals. Among the 730,059 genotyped variants, we excluded 145,238 variants with incorrect annotation, 656 variants with deviation from Hardy-Weinberg equilibrium (HWE) in controls using the statistical threshold of p<10^-6^, 47,286 variants with a Minor Allele Count (MAC) lower than 20, 1,774 variants with a call rate lower than 95%. Finally, 535,105 markers passed the control quality and were used for the imputation. The Imputation was performed with Minimac4 using the 1000 Genomes phase 3 version 5 reference panel [87]. *MARTHA* samples were typed with Illumina Human 610-/ 660W- Quad Beadchip. Quality control procedures of produced genome-wide genotype data in *MARTHA* participants have already been extensively described [88–90]. After further exclusion of individuals with cancer and systemic lupus erythematosus, 1525 participants remained for association testing.

#### Genome Wide Association Study on CFHR5 plasma levels

All SNPs with imputation quality criterion greater than 0.30 and minor allele frequency greater than 0.01 in each participating cohorts were tested for association with CFHR5 plasma levels. Associations were assessed using a linear regression model adjusted for age, sex and study-specific principal components derived from genome-wide genotype data. Results obtained in the different contributing cohorts were then meta-analyzed through a random effect model as implemented in the GWAMA software [91].

## Data Availability

All data produced in the present study are available upon reasonable request to the authors following review of alignment with ethical permits.

## ACKNOWLEDGEMENTS

The study was supported by grants from Stockholm City Council (SLL) to JO (2017-0842/0587, 2020-0346), from Familjen Erling Personssons Stiftelse to MU, Knut och Alice Wallenberg foundation to JO (2020.0182, 2020.0241), from Swedish Heart Lung Foundation to LB (20170759, 20170537), National Research Council (VR) to LB (2019-01493), HelseNord to JO (HNF1544-20). The Human Protein Atlas (HPA) is funded by The Knut and Alice Wallenberg Foundation.

*MARTHA* and *FARIVE* related genetics research programs were funded by the GENMED Laboratory of Excellence on Medical Genomics [ANR-10-LABX-0013], a research program managed by the National Research Agency (ANR) as part of the French Investment for the Future, and supported by the French INvestigation Network on Venous Thrombo-Embolism (*INNOVTE*). *MARTHA* and *FARIVE* genetic data analyses benefit from the technical support of the CBiB computing centre of the University of Bordeaux. GM and D-AT are supported by the EPIDEMIOM-VT Senior Chair from the University of Bordeaux initiative of excellence IdEX. GM benefited from the EUR DPH, a PhD program supported within the framework of the PIA3 (Investment for the future). Project reference 17-EURE-0019. The RETROVE study was supported by grants PI12/00612 and PI15/0026. Genotyping of the *RETROVE* samples was supported by grant PT17/0019, of the PE I+D+i 2013-2016, funded by ISCIII and ERDF.

## Data usage

We used data from the Genotype-Tissue Expression (GTEx) Project (<u>gtexportal.org</u>) [40]. The GTEx project was supported by the Office of the Director of the National Institutes of Health, and by NCI, NHGRI, NHLBI, NIDA, NIMH, and NINDS.

## Figures

Some parts of the figures were created with BioRender.com.

