## Supplemental figures for "Elevated plasma Complement Factor H Regulating Protein 5 is associated with venous thromboembolism and COVID-19 severity"

Figure S1

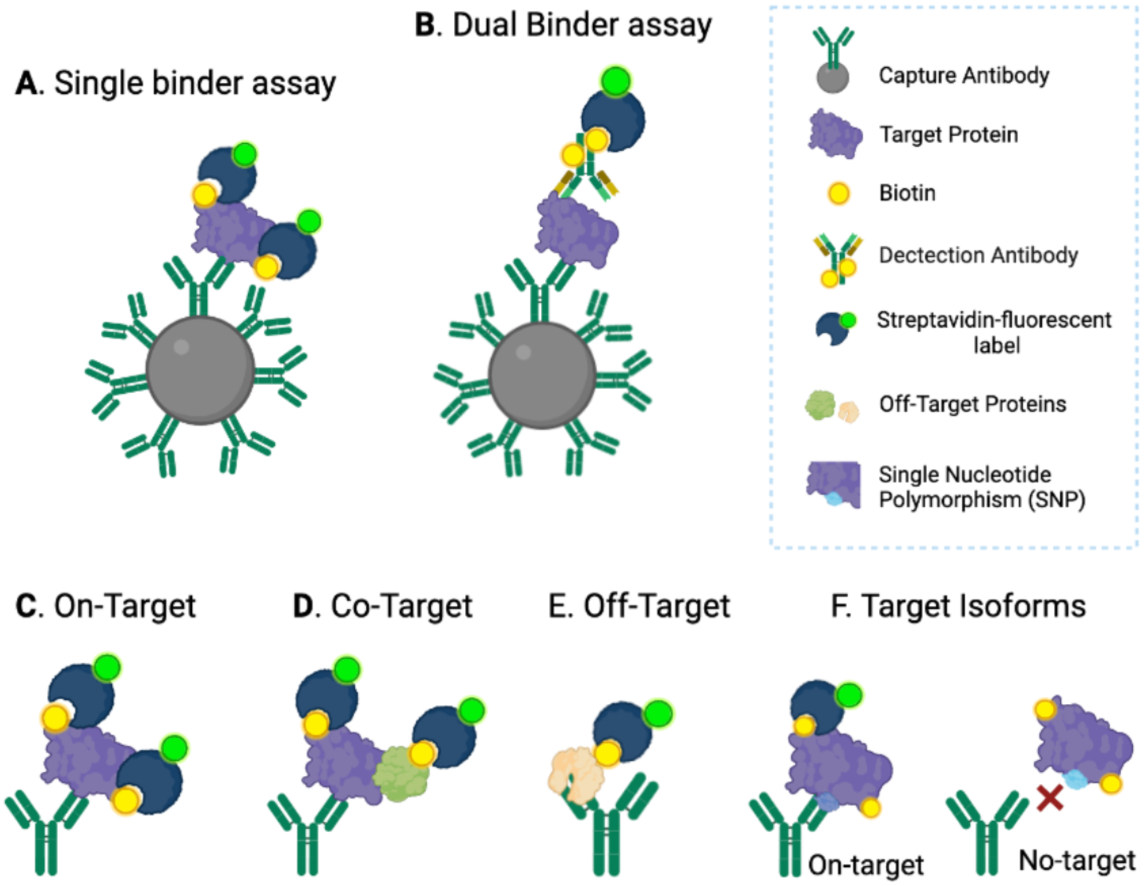


**Figure S1: Antibody based suspension bead assay concepts.** The antibody raised towards intended protein target is covalently coupled to colour-coded micrometre-sized beads. (**A**) In the single binder assay, all proteins within the plasma sample are labelled with biotin before incubation with beads. Biotin-labelled proteins bound to the capture antibody are detected by fluorescent-labelled-streptavidin. The suspension is analysed by a cytometry-based instrument (Luminex), where the colour of the antibody-coupled bead provides the antibody ID, and the mean fluorescence intensity provides a relative measure of the bound target protein corresponding to plasma levels. (**B**) In the dual binder assay captured proteins are unlabelled, and detection of target protein bound to the capture antibody is through a secondary target-specific biotin-labelled detection antibody, followed by fluorescent-labelled-streptavidin addition and analysis on a Luminex instrument. As detection in the single binder assay is based on detection of biotin bound directly to target proteins, the signal can reflect either (**C**) the intended on-target binding, (**D**) co-target binding where the target protein is complexed with non-target protein or (**E**) off-target binding, i.e., binding of a non-target protein**.** (**F**) An amino-acid substitution in the epitope of the target protein can lead to iso-form specific binding.

Figure S2

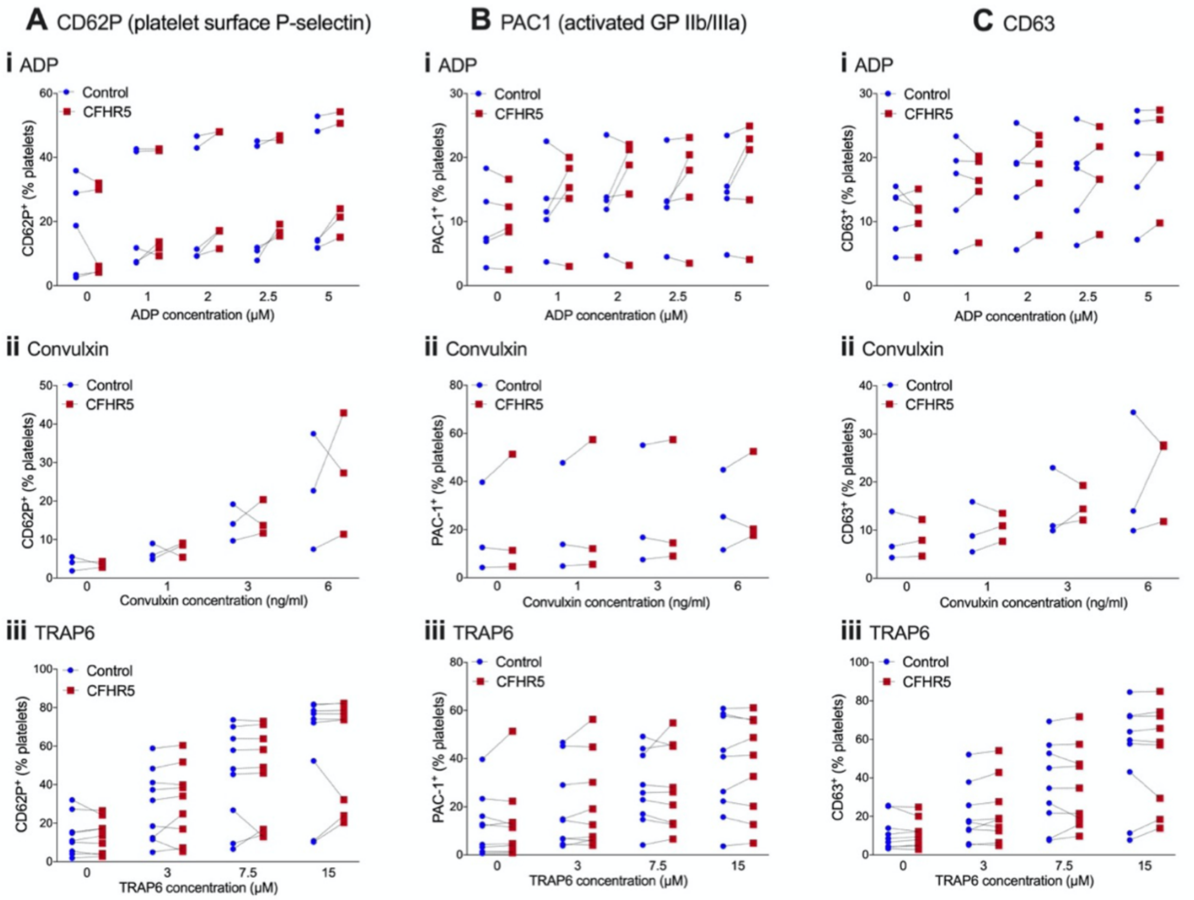


**Figure S2: Recombinant CFHR5 does not potentiate platelet activation of washed platelets.** Platelet activation was measured by surface expression of (**A**) P-selectin, (**B**) activated GP IIb/IIIa or (**C**) CD63, following treatment of washed platelets with different concentrations of (**i**) adenosine diphosphate (ADP) (**ii**) convulxin or (**iii**) TRAP6, following pre-incubation (10 minutes) with 6 µg/ml recombinant CFHR5, or PBS control. Each experiment is represented by an individual point and paired experiments connected by a dotted line.

**Data file Table S1:**

Tab_1: Antibody reagents and target protein IDs from analysis of *VEBIOS ER* discovery study

Tab_2: List of proteins identified in plasma by IC-MS using antibody HPA059937

Tab_3: Association between CFHR5 plasma levels and clinical variables in *VEBIOS ER*

Tab 4: *CFHR5* mRNA co-expression profile in human liver

Tab_5: STRING protein-protein interaction analysis between CFHR5-related transcripts

Tab_6: Relative risk of VTE associated with CFHR5 by thrombosis type and sex

Tab_7: Genetic risk association of VTE

Tab_8: Thrombin generation potential analysis in *MARTHA* and *RETROVE*

**Data file Table S2:**

Tab_1: Clinical characteristics: Extended *VEBIOS Coagulation* sample set

Tab_2: Clinical characteristics: *DFW-VTE* sample set

Tab_3: Clinical characteristics: *FARIVE* sample set

Tab_4: Clinical characteristics: *RETROVE* sample set

Tab_5: Clinical characteristics: *MARTHA* sample sets

Tab_6: Clinical characteristics: *COMMUNITY* study
